# On conducting diary-based social contact surveys to inform transmission models of close-contact infections: a systematic review and expert-informed questionnaire

**DOI:** 10.64898/2026.11.06.26367418

**Authors:** Neilshan Loedy, Giulia Lumia, Laura Di Domenico, Janik Suer, Mark Jit, Alessia Melegaro, Sebastian Funk, Jantien A. Backer, Carlos G. Grijalva, Daniela Paolotti, Joeri Verbiest, Oliver Ratman, Pietro Coletti, Shozen Dan, Vittoria Colizza, Philippe Beutels, Lander Willem, Steven Abrams, Niel Hens

**Author notes:** These authors contributed equally to this work.

## Abstract

**Background:** Diary-based social contact surveys are widely used to parameterise transmission models of close-contact infections. However, variation in study design, contact definitions, and questionnaire content limits comparability across studies. These challenges became particularly evident during the COVID-19 pandemic, when rapid and repeated data collection was urgently needed, often without harmonised instruments. To address these challenges, we aimed to develop an expert-informed questionnaire and update the recommendations on conducting diary-based social contact studies previously proposed by Hoang et al. (2019).

**Methods and findings:** We conducted a systematic review of diary-based social contact surveys published between 31 January 2018 and 25 September 2025 using PubMed and the Web of Science Core Collection, extracting information on study design, survey methodology, contact definitions, and survey items from available questionnaires. Methodological quality and reporting were assessed using an adapted AXIS tool, with results summarised for each criterion. In total, we identified 129 studies across 62 countries, with most studies conducted during the COVID-19 pandemic (59.7% in 2021–2022). Retrospective reporting (86.0%) and non-probability sampling (77.5%) were largely adopted and became increasingly common during the pandemic, while non-paper-based data collection rose sharply from 2021 onwards. Across 36 retrievable questionnaires from general-population surveys, we identified 54 survey items. An online assessment involving 26 experts informed the classification of these items according to their perceived importance, resulting in an expert-informed questionnaire comprising 22 core items. Based on our findings, we updated previous recommendations on sampling, survey methods, contact definitions, behavioural change, essential contact information, and data governance.

**Conclusions:** This study provides an expert-informed questionnaire and updated recommendations to support harmonised social contact data collection, improve comparability across studies, and strengthen preparedness for future public health emergencies.

## 1. Background and rationale

The ways in which people interact influence how infectious diseases spread through populations and how their transmission can be controlled. For pathogens transmitted through close-proximity interactions (e.g., influenza, SARS-CoV-2, and measles), characterising who interacts with whom, in which contexts, for how long, and how frequently those interactions occur can help identify potential transmission pathways and evaluate prevention and control strategies (1,2). Modern transmission models increasingly incorporate heterogeneity in individual interactions and population mixing patterns to improve realism, however the added value of such detail critically depends on availability of empirical data to inform parameterisation and interpretation (3). Social contact surveys provide an important source of such data, which can be used to construct age- and setting-specific contact matrices that inform transmission models (1). Synthetic approaches can provide a scalable alternative where empirical estimates are unavailable (4–7). However, empirical contact data remain essential for characterising population-specific mixing patterns, as well as informing and evaluating these approaches.

Empirical contact data can be collected using several approaches, including proximity sensors, direct observation, digital co-location data, and diary-based social contact surveys (1,8–11). Among these, diary-based surveys have been a commonly used approach for characterising social mixing patterns relevant to transmission modelling, owing to their ability to capture contextual information about interactions, their relative ease of administration, and their applicability across a wide range of settings (12). Within this approach, participants typically report their contacts over a defined reporting period, which may range from a single day to several days or longer depending on the study design. Data collection can follow either a prospective design, where interactions are recorded as they occur, or a retrospective design, in which participants recall their interactions afterwards (13). The criteria used to define a contact may differ across studies, including conversational contacts involving physical presence, direct physical contact, proximity-based encounters, or combinations of these criteria, sometimes with a minimum duration threshold (14). In addition to reporting their contacts, participants are typically asked to provide demographic and contextual information for each contact, which commonly includes the age and sex of the contactee, as well as the setting, frequency, intimacy and duration of the interaction.

A prominent example of a diary-based social contact survey is the POLYMOD study, which collected empirical social contact data from eight European countries that could be used to parameterise infectious disease transmission models (1). Building on this data collection framework, many subsequent diary-based social contact surveys have adopted or adapted the approach across diverse populations and settings. Data from these studies have been extensively used for comparative analyses and scenario modelling, including evaluations of non-pharmaceutical interventions (NPIs) and vaccination strategies (15–21). Although the POLYMOD framework has informed the development of many diary-based social contact surveys, variation remains in their study designs, contact definitions, target populations and subgroups, and data collection methods. These differences might limit comparability across studies and complicate the interpretation of observed contact patterns and their use in transmission models, particularly when comparing populations or assessing changes in mixing patterns over time (22). The COVID-19 pandemic further accentuated these challenges, as the rapid and repeated deployment of contact surveys in response to the pandemic was accompanied by substantial changes in population mixing patterns under NPIs, alongside variation in reporting periods, survey administration approaches and data formats (14,23–26). These experiences illustrate the importance of documenting the epidemiological and policy context in which contact data are collected, alongside harmonising survey methods and questionnaire content to support interpretation and comparison across studies. More broadly, harmonised approaches to survey design and data collection can strengthen the comparability of contact data across populations and over time, supporting their use in transmission modelling and public-health preparedness.

Beyond how surveys are designed and conducted, a related but distinct challenge concerns the content of social contact questionnaires. Although a set of information about the participant, the contacted person, and the interaction between them is commonly collected, the inclusion of additional potentially relevant variables varies across studies (27,28). Recent modelling studies have illustrated the potential value of collecting additional characteristics of contacted persons (e.g., socioeconomic status), which may help characterise heterogeneity in mixing patterns beyond that captured by age and gender and affect estimates of epidemiological outcomes (29,30). Contact diaries could provide a means of collecting such information alongside data on contact events. However, including additional questions increases respondent burden and may compromise data quality (31,32). Determining which survey items are essential, which provide additional value, and which are not justified given the respondent burden, epidemiological relevance and potential effects on data quality remains an open question that has received limited systematic attention in the literature so far.

This study aims to address this gap by developing an expert-informed questionnaire for routine diary-based social contact surveys in the general population and updating the recommendations for social contact studies proposed by Hoang et al. (2019) (27). To achieve these aims, we combined a systematic review of social contact studies published between 2018 and 2025, building on the search strategies of previous reviews conducted prior to the COVID-19 pandemic (27,28), with a structured expert assessment involving researchers in infectious disease modelling, epidemiology, and survey methodology. We used questionnaire content identified in the systematic review to derive potential survey items, which were subsequently assessed by experts and classified as core, secondary, or optional according to their perceived importance for characterising heterogeneity in contact patterns. By integrating evidence from the literature with expert input, this study provides a harmonised questionnaire that aims to better characterise heterogeneity in contact patterns, alongside updated recommendations for diary-based social contact surveys to support more consistent and comparable data collection for transmission models.

## 2. Methodology

### 2.1. Systematic review: search strategy and eligibility criteria

We conducted our systematic review in accordance with the “Preferred Reporting Items for Systematic Reviews and Meta-Analyses” (PRISMA) guidelines. The completed PRISMA checklist, and PRISMA for Abstracts checklist are provided in Supplementary Materials 1 and 2, respectively (33).

#### Search strategy

**PubMed** and **ISI Web of Science (WoS) Core Collection** were searched without language restrictions to identify publications reporting primary studies that collected social contact data. The search strategy was developed based on previous systematic reviews of social contact surveys by Hoang et al. (2019) and Mousa et al. (2021) (27,28), incorporating social contact and mixing-related terms used in these reviews together with terms identifying survey, questionnaire, diary, and panel-based studies. However, unlike Mousa et al. (2021), whose search was restricted to low- and middle-income countries, our search did not impose geographic or income-level restrictions. The following search string was used (see Table S1 for the complete search strings, adapted to the syntax of each database):

(survey*, questionnaire*, diary, diaries, panels*) AND (social contact*, mixing behavio*, mixing pattern*, contact pattern*, contact network*, contact survey*, contact data, contact mixing, contact rate*, contact matri*, contact parameter*, physical contact*, social mixing, mixing matri*, assortative mixing, disassortative mixing, mixing parameter*).

Searches encompassed studies published from 31 January 2018 to 25 September 2025, thereby covering the period after the previous systematic review considered by Hoang et al. (2019) (27).

#### Inclusion criteria

We considered studies eligible if they fulfilled all of the following criteria: (1) the primary focus was on face-to-face contacts between humans, implying the physical presence of at least two individuals during a contact; (2) recording contacts relevant for the transmission of close-contact infections; (3) contacts were recorded using a diary-based approach, either paper-based or electronic; (4) studies reported sufficient methodological detail to allow extraction of survey design characteristics (e.g., contact definition, mode of data collection, sampling, etc.); (5) a full-text version of the manuscript was available in English.

#### Exclusion criteria

We excluded studies that involved at least one of the following: (1) primary focus on human-animal or animal-animal contacts; (2) recording contacts exclusively relevant for sexually transmitted, food-, vector-, or water-borne diseases; (3) using exclusively proximity sensor devices, observational methods, or case-contact investigations to collect contact data; (4) collecting information on contacts without physical presence (e.g., phone, internet/social media contacts) or without the possibility to distinguish them from face-to-face contacts; (5) recording only the frequency or regularity but not the number of contacts over a given time period; (6) studies without a clearly defined recall period for reporting contacts; (7) not being a full-text, peer-reviewed journal article (e.g., meeting abstracts, books, theses, preprints, or other non-peer-reviewed material); (8) secondary analysis of previously published social contact datasets without additional data collection.

#### Data extraction and analysis

Four authors (N.L., G.L., L.D.D., and N.H.) designed a standardised data extraction form prior to extracting information from the included studies (see Supplementary Material 3). All retrieved records were imported into **EndNote X7** for reference management. Bibliographic duplicates were removed using a two-step workflow: automatic identification based on DOI and title matching in EndNote X7, followed by manual verification using title, author, and publication year. For preprint-publication pairs, the peer-reviewed article was retained. Title, abstract, and full-text screening were conducted independently by two reviewers (N.L. and G.L.), with disagreements resolved through consensus or consultation with a third reviewer (L.D.D.). Data extraction for all variables was performed by consensus between at least two reviewers (among N.L., G.L., and L.D.D.).

Extracted information was organised into the following domains: (1) study characteristics, including year of publication, data collection period, and country or region in which the study was conducted; (2) survey methodology, including study setting, study population, sample size, response rate (when available or applicable), availability of incentives, availability of pilot study, sampling design, data collection tools and modes, study design, and reporting period; (3) contact characteristics, in particular contact definitions and type of contact (i.e., physical, conversational, proximity-based, or combinations thereof); (4) availability of survey questionnaire. For the CoMix initiative, which comprises diary-based social contact surveys conducted across more than 20 countries, we extracted data from country-specific analyses when available as published articles. For countries without a separate country-specific publication, information was obtained from published multi-country analyses of the CoMix study (34). In all cases, countries were treated as independent study entries, reflecting distinct data collections, even when multiple countries were reported within a single reference article. As a sensitivity analysis, we repeated the descriptive analyses using initiatives rather than individual survey entries as the unit of analysis (Table S2).

The methodological quality and reporting of included studies were appraised using the AXIS critical appraisal tool for cross-sectional studies (35), adapted to align with our specific study objectives (Supplementary Material 4). Each of the 20 criteria was scored as either 0 or 1, and the proportion of studies meeting each criterion was summarised to identify methodological and reporting aspects that were consistently met or less frequently reported across studies.

### 2.2 Expert assessment of survey items

The questionnaires identified through the systematic review were used to generate a pool of candidate survey questions or measures (hereafter, “survey items”) for expert assessment. We extracted survey items from questionnaires used in surveys conducted in general community settings (see Section 3.3 for the number of questionnaires retrieved), defined as data collected from individuals recruited in the general population, without restriction to a particular institution, workplace, or service setting. Three reviewers (N.L., G.L., and L.D.D.) independently screened the extracted items to identify and exclude items that were specific to COVID-19 contexts (e.g., “Are your encounters less now due to COVID-19?”).

Disagreements regarding inclusion were discussed among reviewers and resolved by consensus. The survey items retained after screening were subsequently reviewed to identify redundancies and improve conceptual clarity. Where multiple studies used closely related questions considered to capture the same information (e.g., “What is your age?” and “What is your date of birth?”), items were merged or rephrased into a single representative item. This process resulted in a consolidated list of 54 survey items included in the expert assessment survey. These were grouped into eight thematic categories: Demographic factors, Socio-Economic (SES) factors, Household composition, Health status, Other (participant) questions, Contact features, Additional contacts (i.e., contacts that could not be recorded individually, typically because of their large number in particular settings), and Information about the survey day.

These 54 survey items were evaluated in a single-round online expert assessment aimed at identifying the items most valuable for routine social contact data collection while accounting for practical considerations related to their implementation. Experts were asked to rate the importance of each item, defined as the perceived value of an item for characterising contact patterns. Importance was used as the primary criterion for item selection, following a similar principle to that used by Thorn et al. (2017) to identify core questionnaire items (36). To complement the importance ratings, experts were also asked whether further research was needed to establish the added value of potentially underexplored items for characterising contact patterns (yes/no). This question helped identify items that experts might consider valuable despite their limited use in existing surveys. Experts also rated the burden (the time and cognitive effort required from participants) and accuracy (the perceived accuracy of their responses) of each item to examine the associations between importance, burden, and accuracy, providing additional insights into the trade-offs involved in questionnaire design (Table S3). Importance, burden, and accuracy were each assessed using three ordinal response categories (1 = Low, 2 = Medium, 3 = High). The expert assessment informed the construction of the proposed expert-informed questionnaire (see Section 2.3).

Experts were instructed to focus on the underlying concept addressed by each survey item, rather than on its exact wording or example responses, when providing their assessment. Mean scores were then calculated across participants for each survey item and thematic category. Pairwise associations between different dimensions were quantified using polychoric correlation coefficient. Accordingly, 95% confidence intervals for the correlation coefficients were estimated using a non-parametric participant-level cluster bootstrap with 10,000 replicates. Furthermore, experts were asked whether any contact location settings identified through the retrieved questionnaires, but not included in the POLYMOD study, should be incorporated into the survey (i.e., other’s home, market/shop, restaurants/bar/café, sports activities, place of worship), and were also invited to provide their overall views on social contact surveys. Prior to deployment, survey questions were pilot tested with a small number of experts and refined based on their feedback. The complete list of survey items and instructions included in the expert assessment is provided in Supplementary Material 5.

In this study, experts were defined as researchers (aged 18 years or older) with professional experience in at least one of the following areas: collection of social contact data, infectious disease epidemiology, or mathematical modelling of infectious disease transmission. Potential participants were purposively identified by reviewing the authorship lists of publications included in the systematic review. Researchers who appeared as authors across multiple included publications were prioritised for invitation, with consideration also given to achieving representation across geographical regions. Additional potential participants were identified through the professional networks of the study authors. The assessment was administered as an online questionnaire via Google Forms, accessible through a link distributed by email. A total of 50 experts were invited, of whom 26 completed the assessment (response rate: 52%). Participation was voluntary, and no incentives were provided.

### 2.3 Expert-informed questionnaire construction

As described in Section 2.2, importance was used as the primary criterion for item selection. In this section, we describe the two-stage procedure used to classify survey items and construct the expert-informed questionnaire. In the first stage, we aimed to identify items that were consistently considered at least moderately important while accounting for variability in expert ratings. Items were therefore retained if the lower bound of the 95% confidence interval for the mean importance score, estimated using a non-parametric bootstrap with 10,000 resamples, was ≥ 2 on the 3-point scale. Items that did not meet this criterion were classified as “optional”. In the second stage, the aim was to distinguish retained items with stronger expert endorsement from those meeting the minimum importance criterion. Items were classified as “core” if their mean importance score was ≥ 2.5, corresponding to an average rating closer to “High” than to “Medium” importance, and as “secondary” otherwise. Among the contact location settings not included in POLYMOD, those endorsed by more than 50% of experts were selected for inclusion in the core questionnaire. Consequently, core items reflect higher perceived importance and serve as the basis of the proposed expert-informed questionnaire; secondary items represent moderate perceived importance and may be used when a broader set of items is required; and optional items reflect lower perceived importance and could be retained for context-specific use. As a sensitivity analysis, the second-stage classification was repeated using the median importance score instead of the mean. Under this approach, retained items were classified as “core” if the median rating was 3 (“High” importance) and as “secondary” otherwise. In addition, the full distribution of expert importance ratings was reported to provide a complementary view of expert agreement.

### 2.4 Updating recommendations

We reassessed the recommendations proposed by Hoang et al. (2019) for conducting diary-based social contact surveys in light of findings from our systematic review and expert assessment (27). For each original recommendation, we compared the findings reported by Hoang et al. (2019) with those from our study to determine whether the recommendation should be retained or updated. Recommendations were retained without modification when our findings did not provide a sufficient basis for change, and were refined, extended, or clarified when supported by new findings. Conceptually overlapping recommendations were consolidated where appropriate, and additional recommendations were developed to address methodological considerations not explicitly covered by the original recommendations. A detailed comparison of the original and updated recommendations, including the supporting evidence from Hoang et al. (2019) and the present study, as well as the status of each recommendation, is provided in Table 2 (27).

**Table 1.**
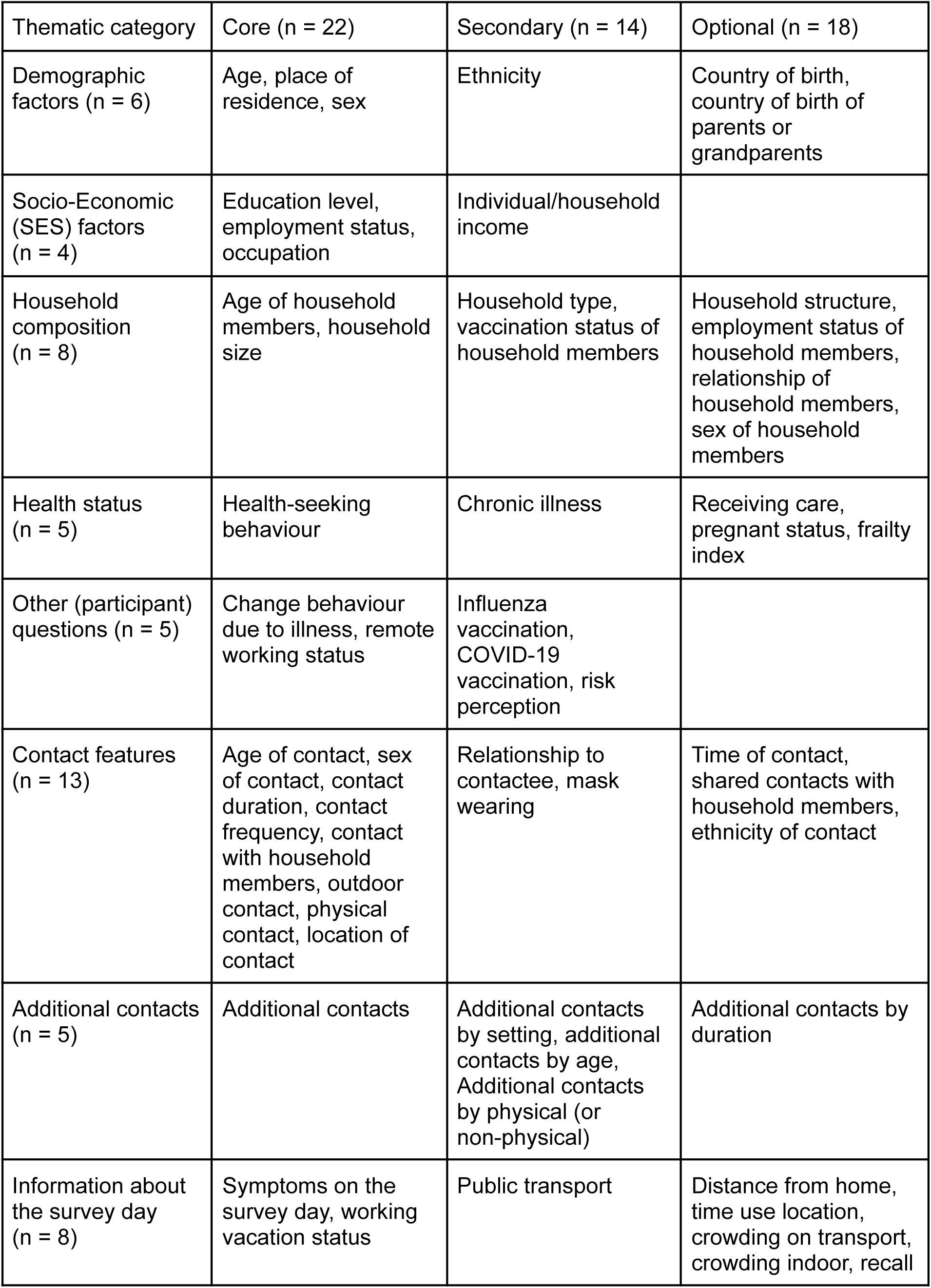
Classification of survey items based on expert-informed questionnaires.

| Thematic category | Core (n = 22) | Secondary (n = 14) | Optional (n = 18) |
|---|---|---|---|
| Demographic factors (n = 6) | Age, place of residence, sex | Ethnicity | Country of birth, country of birth of parents or grandparents |
| Socio-Economic (SES) factors (n = 4) | Education level, employment status, occupation | Individual/household income |  |
| Household composition (n = 8) | Age of household members, household size | Household type, vaccination status of household members | Household structure, employment status of household members, relationship of household members, sex of household members |
| Health status (n = 5) | Health-seeking behaviour | Chronic illness | Receiving care, pregnant status, frailty index |
| Other (participant) questions (n = 5) | Change behaviour due to illness, remote working status | Influenza vaccination, COVID-19 vaccination, risk perception |  |
| Contact features (n = 13) | Age of contact, sex of contact, contact duration, contact frequency, contact with household members, outdoor contact, physical contact, location of contact | Relationship to contactee, mask wearing | Time of contact, shared contacts with household members, ethnicity of contact |
| Additional contacts (n = 5) | Additional contacts | Additional contacts by setting, additional contacts by age, Additional contacts by physical (or non-physical) | Additional contacts by duration |
| Information about the survey day (n = 8) | Symptoms on the survey day, working vacation status | Public transport | Distance from home, time use location, crowding on transport, crowding indoor, recall |

**Table 2.** Comparison of the recommendations proposed by Hoang et al. (2019), the updated recommendations, supporting findings, and recommendation status.

| Subject | Previous recommendations (Hoang et al., (2019)) | Supporting findings from Hoang et al., (2019) and the current study | Updated recommendations | Status |
|---|---|---|---|---|
| 1. Study object | Continue collecting social contact data, particularly in low- and middle-income countries. In high-income countries, surveys can provide further detail on interactions in epidemiologically relevant groups. | - **Hoang et al.:** 64 surveys across 24 countries and five continents, most were conducted in high-income countries, with Europe accounting for 40 surveys.<br>- **Current study:** 129 surveys across 62 countries; South America and Oceania remained underrepresented. | Continue collecting social contact data across diverse populations and geographical settings, particularly in underrepresented regions and populations. | Retained without modification |
| 2. Random sample | Depending on the study objectives, participants should be selected as randomly as possible, so findings can be properly extrapolated to encompass the target population. | - **Hoang et al.:** Convenience sampling was used in 44% of surveys; only seven surveys used random sampling, and two were considered nationally representative. Response rates were reported for 36 of 64 surveys. Only three surveys considered response rates when estimating sample size; 20 used demographic sampling weights to reduce sample bias.<br>- **Current study:** Non-probability sampling was used in 100/129 surveys (77.5%), particularly in 2021 (87.2%) and 2022 (86.8%). Representativeness and response-rate reporting were also among the methodological aspects that least frequently met AXIS appraisal criteria. | Use probability-based sampling where feasible. When this is not possible, consider quota sampling and post-stratification weighting. Clearly document sampling procedures, participant flow, and sample representativeness. | Consolidated and updated |
| 3. Sampling | Describe the sampling procedure in detail, including response rates, participant exclusion, and demographic factors considered when determining sample size. |  |  |  |
| 4. Method | Online and paper diaries both proved to be reliable to measure the overall contact matrix, but are associated with different burdens for the participant, also depending on the age. | - **Hoang et al.:** Both paper and electronic diary formats were used, with reported differences in participant burden depending on survey mode and age.<br>- **Current study:** Non-paper-based tools were used in 92.3% of survey entries in 2021 and 94.7% in 2022. Previous research suggests that survey mode may influence the number of contacts reported (see, e.g., (13,40,41)). | Select the survey mode according to the study objectives, considering its potential effects on participant burden, participation, and contact reporting. Document the mode and distinguish non-applicable responses from missing responses and non-response. | Updated |
| 5. Contact definition | At least two contact types should be included, one aimed to measure more casual contact and one aimed to measure physical contact. | - **Hoang et al.:** Contact definitions varied. Forty of 64 surveys (63%) distinguish physical from nonphysical contacts, but definitions of nonphysical contact differed; 15 surveys used only one contact type.<br>- **Current study:** A combined physical or conversational definition was used in 108 of 129 surveys (83.7%). Proximity-based and other definitions were less frequent, with considerable variation within these categories. | Retain a consistent contact definition: a two-way conversation of at least a few words in the physical presence of another person (within three metres) or direct physical (skin-to-skin) contact. When using this definition, record separately whether each reported contact involved a conversation and whether it involved physical touch. | Updated |
| 6. Prospective versus retrospective | Prospective design should be preferred to retrospective design, since it allows respondents to remember more contact features. | - **Hoang et al.:** Retrospective reporting was used in 29 of 64 surveys (45%), while prospective reporting was used in 26 (41%); four surveys used both approaches.<br>- **Current study:** Retrospective reporting was used in 111 of 129 surveys (86.0%), whereas prospective reporting was used in only 13 (10.1%). | Prospective is preferable over retrospective reporting to reduce recall bias, while considering the recall period, anticipated response rate, and study objectives. | Updated |
| 7. Essential contact information | Age and sex of the contacted person should definitely be included, as well as the duration, the frequency (intimate nature), and the location in which the contact took place. | - **Hoang et al.:** Age and sex of contactees were both recorded in 36/64 surveys (56%); 16 (25%) recorded age only. Location, duration and frequency were recorded in 77%, 67% and 52% of surveys, respectively; all three characteristics were recorded together in 27 surveys (42%).<br>- **Current study:** All five contact characteristics previously recommended by Hoang et al. were classified as core items in the expert assessment. The assessment also identified additional core items, several of which were considered underexplored. | Use the 22 items classified as core in the expert assessment as the basis for a standardised contact questionnaire, adapting it to the study objectives and context. | Updated |
| 8. Scaling information | Information on the size of the possible pool of contactees (like household or school size) proves valuable for testing general assumptions on the scaling of human interactions with the size of the population. | - **Hoang et al.:** Participant household size was associated with the number of contacts in 21 surveys, while four surveys found no association.<br>- **Current study:** Household size and additional contacts were classified as core items. Additional contacts may provide disease-specific information, but was also frequently identified as underexplored. | Collect information on the size of the potential pool of contactees and additional contacts where relevant to the study objectives. | Retained without modification |
| 9. Behavioral change | Possible indicators of behavioral changes (feeling ill) should be included as well, given their large impact on disease spread. | - **Hoang et al:** Five studies found that healthy participants reported more contacts than those feeling ill, while one study did not identify this association.<br>- **Current study:** Behavioural change due to illness, health-seeking behaviour, and symptoms on the survey day were classified as core items. Several of these items were also frequently identified as underexplored. | Include indicators of behavioural change due to illness, together with relevant health-seeking behaviour and symptoms on the survey day, with specific items selected according to the study objectives and respondent burden. | Updated |
| 10. Sharing data | Promote dissemination and reuse of social contact data through standardised formats compatible with the *socialmixr* R package (42). | - **Hoang et al:** Several datasets were available in a unified format compatible with *socialmixr*, facilitating dissemination and reuse.<br>- **Current study:** Substantial variation remained in survey design, data collection and questionnaire characteristics. Standardised data formats and metadata can facilitate interoperability and reuse of the underlying data and derived contact matrices. | Establish reproducible data-processing pipelines and standardised metadata. Share de-identified datasets, derived contact matrices, and the full survey questionnaire, including instructions, in public repositories. | Updated |

### 2.5 Ethical considerations

As the systematic review component of this study was based exclusively on published literature, ethical approval was not required. Ethical approval for the expert assessment was obtained from the Sociaal-Maatschappelijke Ethische Commissie (SMEC) of UHasselt (reference number: REC/SMEC/2025-2026/36).

## 3. Results

### 3.1 Systematic review: study selection and screening

Our search retrieved 2,939 non-duplicate records. Following title-level screening, 246 records were assessed at the abstract level, and 136 proceeded to full-text assessment. Full-text assessment resulted in 84 eligible articles, with six additional articles identified through reference-list searching, resulting in 90 articles included in the review (Figure 1). A retrievable questionnaire was available for 48 (53.3%) of the 90 included articles: 36 (75.0%) from general community settings and 12 (25.0%) from setting-specific surveys. Three included articles described multiple surveys (i.e., (34,37,38)), resulting in 129 surveys conducted across 62 unique countries. Subsequent analyses were conducted at the survey level. Among the surveys, 71 (55.0%) were conducted at the national level. The remaining surveys were limited to specific regions or settings and were not designed to be nationally representative. In particular, 37 (28.7%) survey entries were conducted at a subnational level, focusing on a specific region, city, or town, while 21 (16.3%) targeted specific settings (i.e., educational (n = 8), hospital or healthcare (n = 6), institutional (n = 1), occupational (n = 4), or combined settings (n = 3)).

**Figure 1.**
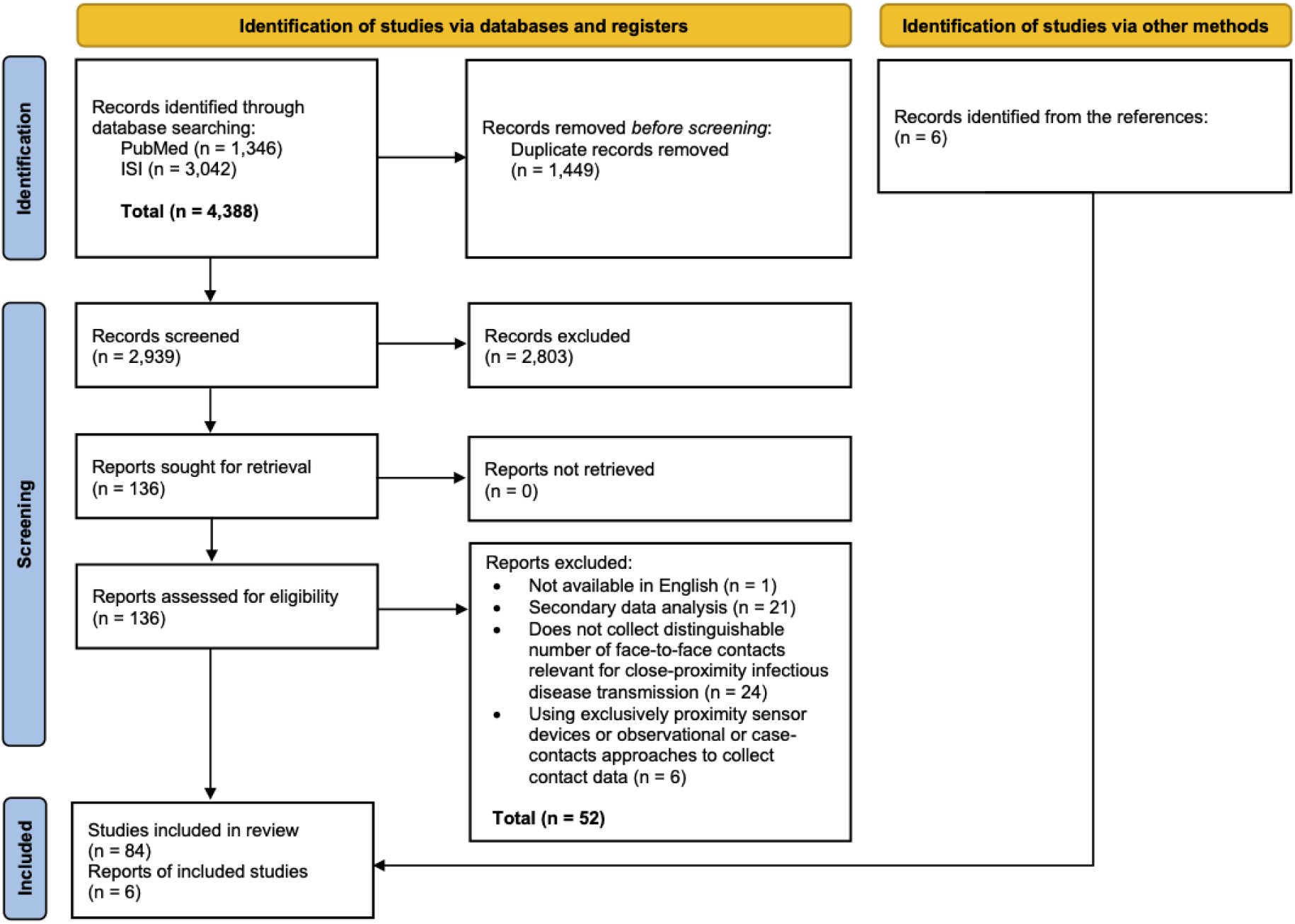
The PRISMA flowchart describing the search process and identification of relevant publications.

The majority of these surveys were conducted in Europe (n = 51; 39.5%), Asia (n = 29; 22.5%), Africa (n = 28; 21.7%), and North America (n = 18; 14.0%). In contrast, relatively few surveys were conducted in South America (n = 1; 0.8%) and Oceania (n = 2; 1.5%) (Figure 2). A small number of countries accounted for a substantial proportion of surveys: the United States (n = 14; 10.9%), China (n = 10; 7.8%), the United Kingdom (n = 8; 6.2%), Belgium (n = 5; 3.9%), the Netherlands (n = 5; 3.9%), and South Korea (n = 5; 3.9%) together represented approximately 35% of the 129 surveys included in the review. The temporal distribution of included surveys was concentrated during the COVID-19 period, with most surveys reported in publications from 2021 (n = 39; 30.2%) and 2022 (n = 38; 29.5%), together accounting for more than half of the surveys included in the review. The reviewed articles met a median of 88.24% of the assessed AXIS criteria (range: 66.67%–100%), indicating generally high adherence to the assessed methodological and reporting criteria (see Supplementary Material 4 for details). Criteria related to clearly stating the aims or objectives, describing the study design, reporting results for the analyses described in the methods, and ensuring that the discussion and conclusions were justified by the results were met by most articles. In contrast, criteria related to the representativeness of participants relative to the target population and reporting of response rates where applicable, were less frequently met.

**Figure 2.**
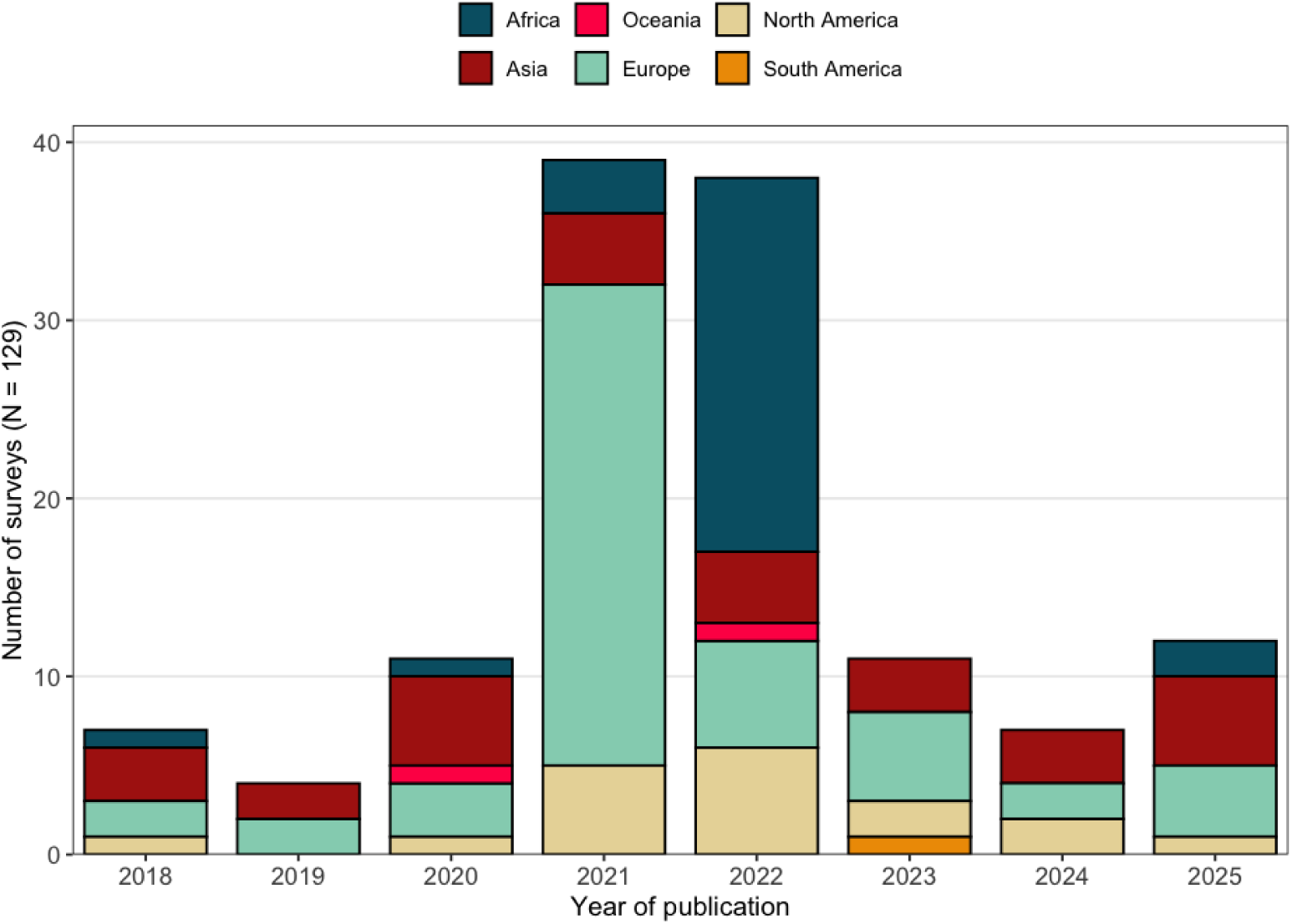
Distribution of the number of surveys by continent and time period.

### 3.2 Systematic review: survey design characteristics

Across the study period, the majority of surveys employed a cross-sectional study design (n = 66; 51.2%), followed by longitudinal (n = 32; 24.8%) and repeated cross-sectional designs (n = 30; 23.3%) (Figure S1). Only one survey (0.8%) combined longitudinal and repeated cross-sectional elements (39). Prior to 2020, longitudinal study designs for social contact surveys were rare, with only one longitudinal survey (0.8%) identified and no repeated cross-sectional surveys reported. In contrast, 15 longitudinal (11.6%) and 4 repeated cross-sectional (3.1%) surveys were identified in 2021, and 3 longitudinal (2.3%) and 20 repeated cross-sectional (15.5%) surveys in 2022, respectively. After 2022, longitudinal and repeated cross-sectional designs became less common, as cross-sectional designs again accounted for the majority of surveys identified in our search. With respect to reporting approach, the majority of surveys relied on retrospective reporting of contacts (111 surveys; 86.0%). Prospective data collection was used in 13 surveys (10.1%), while 5 surveys (3.9%) employed a combined retrospective-prospective approach.

With respect to survey mode, most surveys used self-administered designs (n = 79; 61.2%). Interviewer-administered approaches were less common, including 28 phone-based (21.7%) and 14 face-to-face interviews (10.9%), while 8 (6.2%) employed mixed survey modes. Survey mode and data collection tools varied by publication year: self-administered surveys dominated in 2021 (n = 31/39; 79.5%), whereas phone interviews were more frequently reported in 2022 (n = 19/38; 50.0%). They also varied geographically: self-administered surveys accounted for 47 of 51 surveys in Europe (92.2%) and 17 of 18 in North America (94.4%), whereas phone interviews accounted for 20 of 28 surveys in Africa (71.4%). Survey modes were more heterogeneous in Asia, where self-administered surveys accounted for 14 of 29 surveys (48.3%), followed by face-to-face interviews (n = 6/29; 20.7%), phone interviews (n = 5/29; 17.2%), and mixed modes (n = 4/29; 13.8%). Non-paper-based tools were used most often in 2021 (n = 36/39; 92.3%) and 2022 (n = 36/38; 94.7%), while prior to 2020 paper-based tools were more common (n = 7/11; 63.6%) (Figure S2). Sampling designs were predominantly non-probabilistic (n = 100; 77.5%), while probability-based sampling was reported in 28 surveys (21.7%) and one survey (0.8%) combined both probability and non-probability approaches. Non-probability sampling was most frequently reported in 2021 (n = 34/39; 87.2%) and 2022 (n = 33/38; 86.8%). Most surveys used one-stage sampling (n = 101; 78.3%), with multi-stage sampling reported in 27 surveys (20.9%) and one study (0.8%) combining one- and multi-stage elements.

Definitions of a contact varied across surveys. Most surveys defined contacts as either physical or conversational interactions (n = 108; 83.7%), while smaller proportions used proximity-based definitions (n = 7; 5.4%), combined physical, conversational, and proximity-based definitions (n = 7; 5.4%), conversational contacts only (n = 4; 3.1%), physical contacts only (n = 2; 1.6%), or physical or proximity-based contacts (n = 1; 0.8%). Note that heterogeneity exists within these contact categories (e.g., distance thresholds, minimum duration or number of words exchanged, and whether physical contact was required). We refer to the Supplementary Material 3 for the original contact definitions used in each study. In terms of contact-information level, 120 surveys (93.0%) collected contact data at the individual level, while 9 surveys (7.0%) adopted a household-level reporting structure. Reporting of pilot testing and participant incentives was infrequent, with only 6 surveys (4.6%) and 9 surveys (7.0%) explicitly reporting these, respectively.

### 3.3 Systematic review to expert assessment: survey item identification and consolidation

The extraction and consolidation process yielded 54 survey items from the 36 surveys conducted in general community settings for which questionnaires were retrievable (Section 3.1). The frequency with which these items were captured across surveys varied considerably. Participant age was captured in all surveys, followed by participant sex (33/36, 91.7%), household size, contact age, contact location, and whether contacts involved physical (skin-to-skin) contact (32/36, 88.9% each). Seven items were each reported in only one of the 36 surveys (1/36, 2.8%), including participant characteristics such as remote working status, receiving care, frailty index, ethnicity, and country of birth of participants’ parents or grandparents, as well as contacts in common between household members and an indicator of public transport crowding. Together with the exclusion of COVID-19-specific survey items described in Section 2.2, these findings formed the basis for the final set of items taken forward for expert evaluation. Supplementary Material 6 provides the frequency with which each survey item appeared across the retrieved questionnaires, the source questionnaires from which each survey item was extracted, and its final core, secondary, or optional classification.

### 3.4 Expert assessment: survey item ratings and classification

The expert cohort comprised 26 individuals across five continents (Figure S3). The majority of these experts were affiliated with European institutions (53.8%), followed by North America (23.1%), Asia (15.4%), Africa (3.8%), and Oceania (3.8%). Across survey items, polychoric correlations between item-level ratings given by these experts indicated weak yet significant negative associations between importance and burden (ρ = −0.2; 95% CI: -0.36–-0.17, Figure 3), and positive associations between importance and accuracy (ρ = 0.28; 95% CI: 0.16–0.40, Figure S4). A stronger inverse association was observed between burden and accuracy (ρ = −0.72; 95% CI: -0.79– -0.65, Figure S5), indicating that items perceived as more burdensome were also more likely to be viewed as yielding less accurate responses.

**Figure 3.**
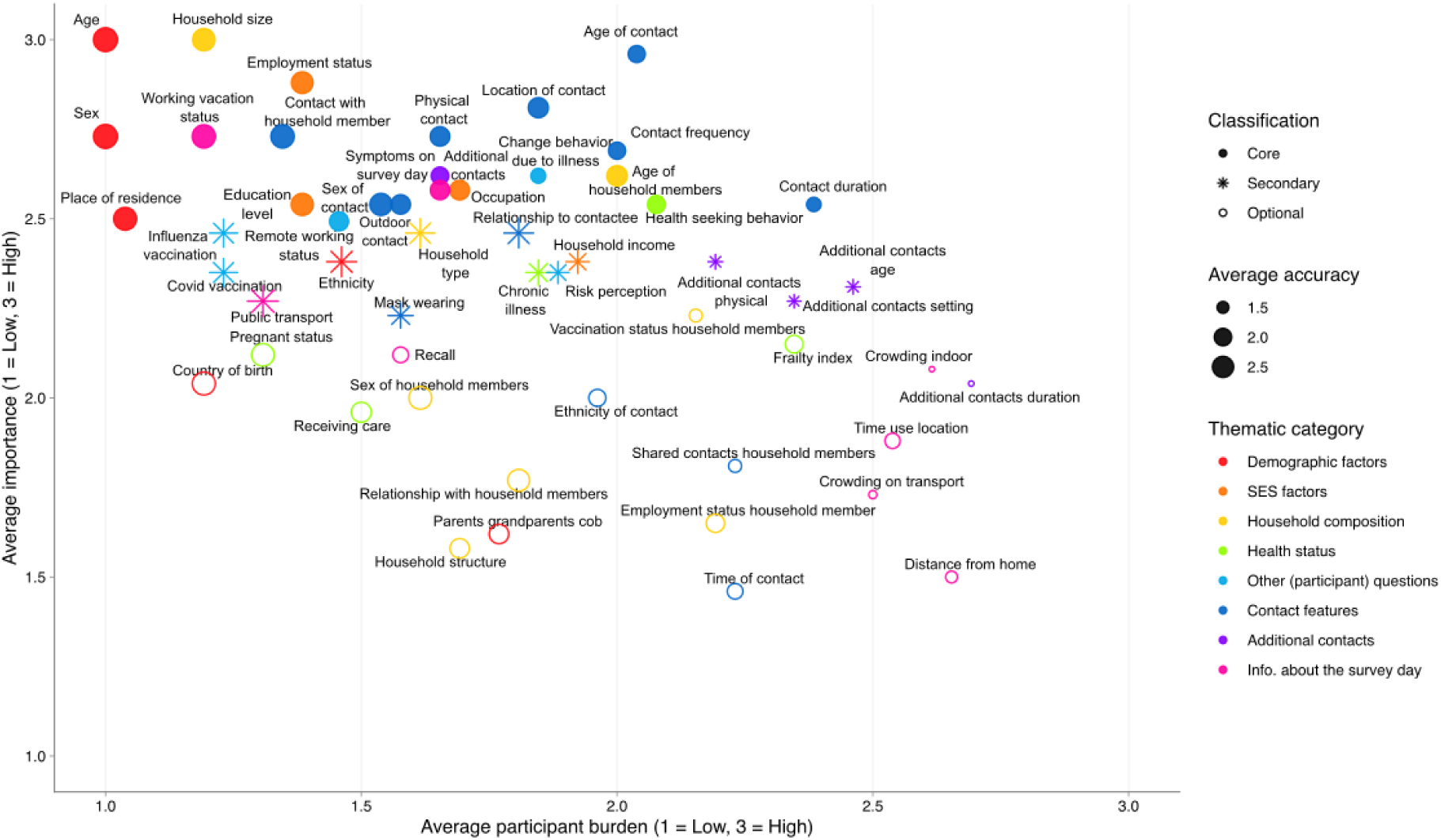
Expert assessment matrix mapped by the average participant burden and average importance, sized proportionally to their average reporting accuracy.

Higher average importance scores were observed among the thematic categories for SES factors (2.60; 95% CI: 2.48–2.70), Other (participant) questions (2.45; 95% CI: 2.35–2.55), Contact features (2.42; 95% CI: 2.35–2.50), and Demographic factors (2.38; 95% CI: 2.26–2.49). Survey items within the Demographic factors and Household composition categories were generally perceived as having high accuracy and low respondent burden (Table S4). In contrast, survey items related to Additional contacts and Information on the survey day categories were perceived as more burdensome and less accurate. Other survey items requiring information about other individuals (e.g., household members’ vaccination status or participants’ parental country of birth) were generally perceived as more burdensome and potentially less accurate than survey items concerning participants’ own characteristics. Although demographic factors had the lowest average proportion of experts identifying survey items as underexplored (39.1%, 95% CI: 31.4–46.8%), several characteristics within this category were still considered to warrant further research, including participants’ ethnicity (69.2%, 95% CI: 50.0–84.6%), household income (61.5%, 95% CI: 42.3–80.8%), and participants’ country of birth and occupation (both 53.8%, 95% CI: 34.6–73.1%) (Figure S6). Similarly, within Contact features (40.2, 95% CI: 34.9–45.6%), outdoor contacts (i.e., information on whether contacts occurred indoors or outdoors) (65.4, 95% CI: 46.2–84.6%) and contact frequency (53.8, 95% CI: 34.6–73.1%) were frequently flagged as underexplored. Health status (65.4%) and Other (participant) questions (66.9%) exhibited the highest proportions of survey items flagged as needing further research. When experts were asked to assess additional contact settings not included in POLYMOD for potential inclusion in social contact surveys, restaurants, bars and cafés, and other people’s homes received the highest endorsement (at 65.4% each), and were therefore included as additional contact settings in the core questionnaire. Endorsement was lower for places of worship (50.0%), markets or shops (46.2%), and sports-related settings (34.6%).

Applying the predefined classification procedure resulted in 22 core, 14 secondary, and 18 optional items (Figure 3). The 22 core items formed the proposed expert-informed social contact questionnaire, which is provided in Supplementary Material 7. Details on the underlying scores used to classify each item into the three categories are provided in Figures S7–S9. Among the 22 core survey items (Table 1), participant age and household size received the strongest endorsement, with unanimous agreement among experts that both were highly important (Figure S10). Other highly endorsed survey items included the age of the contactee (average importance: 2.96; 95% CI: 2.88–3.00), participants’ employment status (2.88; 95% CI: 2.69–3.00), and contact location (2.81; 95% CI: 2.62–2.96). These were rated as highly important by 96.2%, 92.3%, and 84.6% of experts, respectively. Several additional contact-level characteristics, including whether the contact was a household member (80.8%), whether physical (skin-to-skin) contact occurred (76.9%), and participants’ working or vacation status on the survey day (76.9%), also received high levels of endorsement, each with a mean importance score of 2.73 (95% CI: 2.50–2.92). Comparatively weaker endorsement was observed for participants’ place of residence and remote working status (both with an average importance score of 2.50 (95% CI: 2.23–2.73)), although both remained classified as core, with just over half of experts rating these items as highly important. As a sensitivity analysis, the second-stage classification was repeated using the median importance score instead of the mean. This resulted in two items (household type and relationship to contactee) being reclassified from secondary to core items, while all remaining classifications were unchanged.

Survey items relating to SES factors and Other (participant) questions were predominantly classified as core or secondary, indicating that these characteristics were generally considered relevant for inclusion in social contact surveys. Although core items were, on average, less frequently flagged as underexplored (39.9%, 95% CI: 30.8–48.8%) than items classified as secondary (58.8%, 95% CI: 52.7–64.5%) or optional (56.6%, 95% CI: 52.1–61.1%), several core items were nevertheless frequently identified as requiring further research. For example, participants’ remote working status (80.8%, 95% CI: 65.4–96.2%), change in behaviour due to illness (73.1%, 95% CI: 53.8–88.5%), health-seeking behaviour (69.2%, 95% CI: 50–84.6%), and outdoor contacts (65.4%, 95% CI: 46.2–84.6%) were frequently flagged as underexplored despite their classification as core items. In contrast, participant age was not flagged as underexplored by any experts, while contact with household members (3.8%, 95% CI: 0–11.5%) and contact age (11.5%, 95% CI: 0–23.1%) were also flagged as underexplored by few experts.

For secondary items, perceived need for further research was particularly high for vaccination status of household members (76.9%, 95% CI: 61.5–92.3%), chronic illness (73.1%, 95% CI: 53.8–88.5%), ethnicity (69.2%, 95% CI: 50–84.6%), and risk perception of the participants and additional contact by setting (65.4%, 95% CI: 46.2–80.8% each). Within optional items, the proportion of experts who identified a need for further research ranged from 30.8% (95% CI: 15.4–50%) for sex of household members to 80.8% (95% CI: 65.4–96.2%) for indoor crowding. Other items frequently identified as requiring further research included participants’ frailty index (69.2%, 95% CI: 50.0–84.6%) and shared contacts between household members (65.4%, 95% CI: 46.2–84.6%).

### 3.5 General recommendations

Of the 10 recommendations proposed by Hoang et al. (2019) (27), some were retained without modification, while others were consolidated or updated based on findings from the systematic review and expert assessment. Table 2 summarises the original recommendations, the supporting evidence from this study, and the resulting changes. The following sections discuss the practical implications of these recommendations for the design, implementation, and dissemination of diary-based social contact surveys.

#### Sampling strategy

Our review showed that non-probability sampling predominated across studies, encompassing approaches such as convenience sampling, opt-in panels, purposive sampling, quota sampling, and snowball sampling. Where feasible, we recommend using probability-based sampling to reduce the risk of selection bias. When this is not possible, strategies such as quota sampling and post-stratification weighting can be considered to improve the alignment between the achieved sample and the target population (or the intended study population for setting-specific surveys) across key observable characteristics. Regardless of the sampling approach, the sampling strategy should be clearly documented, and participants should be selected to reflect the intended study population as closely as possible. Quota attainment, participant flow, and comparisons between the achieved sample and the target population should also be reported to facilitate assessment of potential selection bias.

#### Method

We recommend selecting the survey mode according to the study objectives and considering its potential effects on participation, respondent burden, and contact reporting. The mode of data collection should be clearly documented to facilitate comparisons across studies. Across survey modes, non-applicable responses should be distinguished from missing responses and non-response, and documented accordingly.

#### Essential contact information

Our expert assessment supports retaining the age and sex of the contacted person, as well as the duration, frequency, and location of each contact, as core questionnaire items. These characteristics were also frequently included in the retrieved questionnaires. We further recommend collecting information on physical (skin-to-skin) touch, household contacts, and contact settings not included in POLYMOD, particularly restaurants, bars and cafés, and other people’s homes. Indoor versus outdoor contact and additional contacts were also classified as core items, with a need for further research to fully establish their influence on social contact patterns and disease spread.

#### Contact definition

Our review found that most study entries used a single combined definition of physical or conversational contact, whereas proximity-based and other combined definitions were uncommon. To support comparability across studies, we recommend retaining a consistent contact definition: either a two-way conversation of at least a few words in the physical presence of another person (within three metres) or direct physical (skin-to-skin) contact. When using this definition, we also recommend distinguishing between contacts involving (skin-to-skin) touch and those involving conversation only. Additional contact information, such as whether the contactee is a household member, the relationship to the contactee, and the environmental setting (e.g., indoor versus outdoor), can be included according to the pathogen of interest and the study objectives. Contacts involving larger groups or settings can be recorded separately where relevant to study objectives.

#### Behavioural change

The inclusion of indicators of behavioural change due to illness remains relatively underexplored in contact survey research. Our findings support collecting information on symptoms on the survey day, behavioural changes due to illness, and health-seeking behaviour, all of which were identified as core items for contextualising changes in contact behaviour. We therefore recommend including these indicators, with specific items selected according to the study objectives and respondent burden.

#### Data governance and dissemination

Reproducible data-processing pipelines, clear data dictionaries documenting variable definitions and coding schemes, and standardised metadata should be established from the outset, in accordance with the FAIR (Findable, Accessible, Interoperable, and Reusable) principles. We also recommend making the full survey questionnaire, including question wording, response options, and relevant instructions, available alongside the data to support transparent interpretation and reuse. Where ethically and legally permissible, we recommend depositing de-identified participant- and contact-level datasets, preserving linkage across diary days, alongside derived contact matrices in public repositories such as socialcontactdata.org and the social contact data community on Zenodo. To facilitate reuse and interoperability, datasets should follow common data formats and metadata standards, such as those developed within SOCRATES (43). Retaining these records alongside derived contact matrices preserves information on individual-level variation and allows datasets to be reanalysed using alternative age groupings, weighting schemes, and assumptions about missing data. Adopting common standards can further facilitate interoperability with analytical tools, including *socialmixr* and *contactsurveys*, thereby improving the long-term scientific value and reusability of social contact data (44,45).

## 4. Discussion

Diary-based social contact surveys remain one of the most widely used and practical approaches for collecting individual-level contact data to parameterise transmission models of close-contact infections. The widespread adoption of POLYMOD-type questionnaires has provided continuity across many studies and served as an important basis for cross-study comparison. However, the increasing number of studies carried out in recent years introduced considerable variations in questionnaire content, item construction and study design, demonstrating the need for standardised instruments and clearer guidance.

Updating recommendations therefore requires balancing established core elements with additional characteristics that may warrant greater attention in future surveys. Drawing on a systematic review of 129 surveys across 62 countries and a structured expert assessment, this study proposes a classification of survey items into core, secondary, and optional categories, which informed the development of an expert-informed questionnaire for diary-based social contact surveys in the general population, with the aim to improve cross-study comparability and support data-sharing initiatives such as SOCRATES and SOCRATES-CoMix (43,46). The present work focuses specifically on questionnaire content and study design guidance. Complementary methodological considerations, including sample size determination and reporting approach, are addressed elsewhere (12,13,47,48).

The proposed expert-informed questionnaire comprises 22 core items derived from the expert assessment and retains most of the information captured by the POLYMOD questionnaire (1), while incorporating new questions. In terms of item count, the proposed core questionnaire falls between POLYMOD (15 items) and CoMix (30 items), providing context for its scope relative to established social contact questionnaires (1,34). The novel core items cover health-seeking behaviour, behavioural change due to illness, symptoms on the survey day, remote working status of the participants, and whether contacts occurred indoors or outdoors. Health-seeking behaviour provides insight into how participants perceive and respond to illness, whereas reported behavioural changes due to illness capture modifications in social activity, such as reducing contacts or staying at home, in response to symptoms or awareness of infection (49). Studies have shown that individuals having for example influenza-like symptoms or underlying frailty generally report fewer contacts. Consequently, contact matrices derived predominantly from healthier participants may not fully represent contact behaviour in the broader population, potentially biasing parameter estimates and epidemic projections (41,50–52). Recording symptoms and illness-related behavioural changes can help further stratify contact patterns and examine their implications for epidemic modelling. In practice, however, capturing this subpopulation is challenging as their willingness to participate may decrease when feeling unwell (41,53). Working and vacation status can also help contextualise reported contact patterns, given substantial variation in daily mixing behaviour across occupational schedules and holiday periods. The inclusion of remote working status is particularly relevant following the COVID-19 pandemic (54), as the widespread and unequal adoption of remote work has altered individuals’ daily contact patterns and social mixing in ways that vary by gender, caregiving responsibilities, and socioeconomic status (55). The setting in which contacts occur is also relevant to understanding transmission. Evidence from SARS-CoV-2 studies suggests that transmission occurs predominantly indoors, although substantial heterogeneity in study quality and definitions of outdoor settings limits conclusions about the relative risk of transmission indoors versus outdoors (56,57). The expert assessment also identified these novel core items as underexplored, signalling the need for further research into how contextual and behavioural factors shape social mixing patterns. In line with the expert assessment, restaurants, bars, and cafés may present settings conducive to respiratory pathogen transmission, attributed to potential overcrowding, cross-infection risk from staff, and reduced ventilation (58). Infection risk for respiratory pathogens also increases with proximity and duration of contact (59), making private residences a relevant setting to capture. Reflecting both this evidence base and the results of our expert assessment, restaurants/bars/cafés and others’ homes were included as explicit location categories in the contact diary.

Ethnicity was identified as a relatively underexplored domain in social contact surveys, and classified as a secondary item. Recent emerging evidence shows that ethnic, migration-related, and socioeconomic characteristics may capture dimensions of contact pattern heterogeneity beyond age. Studies in the United Kingdom and United States have shown that ethnic and socioeconomic characteristics can capture important dimensions of contact pattern heterogeneity beyond age (60–62), while research among migrant workers in Thailand similarly identified clustered contact patterns varying across migrant backgrounds and occupation type (63). Notably, the definition and classification of ethnicity vary across settings: studies in Western high-income settings typically report broad racial categories (e.g., White, Black, Asian) (34,60,64–66), whereas studies from Asian and African contexts more commonly use locally defined ethnic group classifications (37,67–69). Respondent nationality was included in POLYMOD, and could serve as a crude proxy for population ethnic heterogeneity, although the two concepts are not completely equivalent. Legal and institutional contexts may also affect the feasibility and preferred approach to collecting these data, leading some studies to use proxy measures such as country of birth, nationality at birth, or parents’ country of birth (70). These differences in how characteristics are defined and classified emphasise the importance of clearly documenting the variables used to facilitate comparison across surveys.

Beyond ethnicity, socioeconomic characteristics of the participants, such as household income and occupation, represent additional underexplored dimensions of contact pattern heterogeneity. This is particularly relevant given that socioeconomic factors received the highest average importance score among all thematic categories in our expert assessment. Participants’ socioeconomic circumstances may shape their health outcomes and constrain their ability to reduce contacts during periods of non-pharmaceutical interventions (NPIs), suggesting the need to consider heterogeneity beyond age in infectious disease models (61). Recording these characteristics could therefore help characterise differences in contact patterns that may be overlooked by traditional age-stratified models. Although the potential relevance of socioeconomic heterogeneity for contact patterns and transmission modelling has received increasing attention, direct information on contactees’ socioeconomic characteristics remains limited (29,30,71). Obtaining such data is, in fact, challenging, as participants may have limited knowledge of the income or occupation of less familiar contacts. As such, whether more readily reported indicators can capture this heterogeneity remains an open question.

Alongside variation in the survey questions, studies also differ in how they define a contact (14,28,43). Such variation may limit comparability across studies. It is also important to recognise that a reported contact does not necessarily correspond to an exposure event, and infection may occur through multiple routes (48). These considerations support the use of a harmonised contact definition, as proposed in Section 3.5, together with the systematic recording of contact characteristics, including whether physical contact occurred, the duration of the interaction. The specific characteristics collected may be tailored to the study objectives and pathogen of interest. Variation may also arise from the data collection method itself, as different methods capture distinct aspects of interaction (8–11). This should be considered when comparing or combining contact data, as methodological differences can carry through to empirical and synthetic contact matrices and ultimately influence the parameterisation of transmission models (4–6). Our expert assessment further supported collecting these additional contact characteristics, which have been commonly included in large-scale surveys since POLYMOD (1,14). However, the items related to additional contacts received lower ratings for reporting accuracy despite being considered important for understanding transmission dynamics. Survey design should therefore balance the potential epidemiological value of an item against the burden and accuracy associated with obtaining the information.

Beyond questionnaire content, our review also illustrates broader methodological challenges in collecting social contact data. Careful questionnaire development and pre-testing can help identify measurement and practical issues before full-scale implementation, particularly in underexplored settings or populations. However, pilot or pre-test studies were explicitly reported in only 6 of 129 study entries (4.6%), indicating limited reporting of these practices rather than necessarily limited use. Preparing and evaluating survey methods in advance may also support timely data collection during future public health emergencies. Rapid deployment and repeated data collection during public health emergencies can help track changes in contact behaviour, but also introduce challenges related to sampling, reporting, and follow-up (72). In longitudinal studies, repeated participation may contribute to attrition and respondent fatigue, as well as changes in reporting over time. Changes in reporting may also arise from panel conditioning, whereby participation in earlier survey waves affects subsequent behaviour or responses. Statistical approaches to account for fatigue-related under-reporting have been proposed (31,32), although panel conditioning effects remain comparatively less addressed in the social contact survey literature. A focused set of indicators measured consistently across waves may facilitate comparisons while limiting respondent burden. Clear instructions and reminders may also help maintain participation, while recruiting participants at different time points, where appropriate, may help assess the effects of repeated participation (34). Measures of behavioural change can further help contextualise observed contact patterns in relation to illness and public health restrictions.

One important design choice is whether contacts are recorded prospectively or retrospectively. Retrospective reporting was common in our review (86.0% of study entries), compared with 45% in Hoang et al. (2019) (27), despite the methodological preference for prospective reporting to minimise recall bias (13). Previous research comparing the two approaches highlights a trade-off between recall bias and response, with prospective reporting reducing recall bias and retrospective reporting potentially achieving higher response rates. For short recall periods, differences in reported contacts were small, and the potential higher response rate of retrospective reporting was considered to outweigh the disadvantage of slight underreporting (13). The choice between prospective and retrospective reporting should therefore consider the recall period, anticipated response rate, and study objectives. Survey mode and sampling strategy also require careful consideration, as they may affect participation and contact reporting. Digital survey modes may affect participation differentially across respondent characteristics, including age-related differences in mode preferences (40,41). They can also facilitate filtered or conditional questions that reduce respondent burden by presenting items only when relevant; however, responses that are not applicable should be clearly distinguished from missing responses or non-response and documented accordingly (73,74). Where face-to-face fieldwork is impractical, digital surveys may offer a practical alternative, although their suitability should be considered in relation to the target population and study objectives. Together, these considerations underscore the need to align survey and recruitment approaches with study objectives and to clearly document procedures and response patterns to support interpretation and comparison of contact data.

Recent comparisons of empirical and mobility-driven synthetic contact matrices during the COVID-19 pandemic have shown particularly pronounced discrepancies in contact estimates among children and adolescents when schools were open (7). Although these discrepancies may have multiple explanations, they exhibit the uncertainty surrounding how well different approaches capture children’s contacts, particularly in school settings. One methodological consideration is who reports children’s contacts. Depending on the child’s age and ability to report their own interactions, contact diaries may be completed by the child, a parent or caregiver, or with adult assistance. Evidence from other areas of health research suggests that agreement between parent-proxy and child reports varies according to the type of information reported, with better agreement generally observed for more directly observable measures (75). In social contact studies, approaches include age-specific diaries with proxy reporting for younger children, as well as tailored instructions and questionnaires for proxies and teachers to support reporting of contacts in specific settings, such as schools (e.g., (76,77)). Further research is warranted to establish whether, and under what circumstances, these data collection procedures and survey design choices influence the number and information of contacts reported for children and adolescents.

Although social contact studies from low- and middle-income countries (LMICs) remain underrepresented relative to those from high-income countries (HICs), their geographic coverage has increased markedly. Hoang et al. (2019) and Mousa et al. (2021) identified contact studies from 9 and 19 LMICs in studies published up to January 2018 and May 2020, respectively, compared with 48 LMICs in our updated review (27,28,78). This expansion in the geographic coverage of the literature, however, was not mirrored in the composition of our expert panel, which may have limited the representation of priorities specific to LMIC settings in the expert assessment (79). For example, the number of rooms in a household has been collected in several LMIC-based contact studies as a proxy for household crowding and the capacity to isolate during outbreaks (37,80–84). Such information may be particularly relevant in settings where large or multigenerational households are common and opportunities for within-household isolation are limited (28). More broadly, the relevance of ethnicity and socioeconomic factors is highly context-dependent, varying by disease, population, and setting. The questionnaire and guidelines proposed here therefore provide a common starting point that can be adapted to local research priorities, specific research goals, epidemic contexts, intervention requirements, and the cultural and linguistic characteristics of the study setting, potentially lowering barriers to initiating social contact studies in LMICs and other underrepresented settings.

Recommendations informed by systematic review and expert assessment, such as those proposed here, can support more standardised social contact data collection and improve comparability across settings. Their impact, however, depends on robust data governance and sharing frameworks that allow data to be integrated and reused across studies and surveillance systems (85). The COVID-19 pandemic further accentuated that timely cross-national data sharing was critical for epidemic modelling, while differences in variable definitions and data structures complicated the interpretation of model outputs. It also illustrated the value of establishing sustainable data collection systems before an emergency, so that standardised data can be collected and shared globally when needed (25). Preparedness requires complementary resources, including pathogen- and context-specific extensions, generic ethics and consent templates, scalable sampling strategies, and the supporting data standards and sharing infrastructure (25,86). Research groups should also establish clear and reproducible data processing pipelines, from raw data to final output, with data quality checks. Ideally, these pipelines should be integrated within institutional data platforms, where data from past and future studies can be stored permanently, with a managed computing environment, a version control system and a catalogue linking datasets to related publications. The processed data can then be shared more widely using global initiatives. Initiatives such as *socialmixr* and *contactsurveys*, SOCRATES, SOCRATES-CoMix, and broader efforts such as socialcontactdata.org illustrate how coordinated infrastructures can support harmonised data collection and enable rapid multi-country analyses (43–46). The FAIR principles provide a practical way to support this, by ensuring that data are findable, accessible, interoperable, and reusable (87). More broadly, the questionnaire and guidelines developed here may also provide a practical starting point for new social contact studies, allowing researchers to adapt a common baseline to specific populations, settings, or research questions without having to develop the core survey infrastructure from scratch.

Several caveats should be noted. First, experts were asked to examine question items independently. As a result, items capturing overlapping information may have been evaluated separately rather than as part of a coherent questionnaire. Future assessment could incorporate prioritisation methods such as best-worst scaling, which ask experts to identify the most and least important items within small subsets rather than rating each independently (88). Alternatively, evaluating predefined questionnaire modules as a whole may better capture trade-offs between item importance, respondent burden, perceived accuracy, and overall questionnaire length. These considerations also inform how the resulting questionnaire should be interpreted: it represents one possible synthesis of the evidence identified through the review and expert assessment. As item classification was based primarily on perceived importance, the resulting priorities may favour items that are already widely recognised in social contact research over less established items whose potential value remains uncertain. In addition, some experts had authored publications included in the review. Their familiarity with particular survey approaches may have influenced their ratings, potentially favouring items commonly used in existing studies. Nevertheless, the sensitivity analyses show that the item classifications are robust. We also provide the extracted items separately, allowing researchers to select those best suited to their study objectives and context. Furthermore, the expert assessment was conducted in a single round, without an opportunity for experts to reconsider their ratings in light of group feedback. The results therefore reflect individual expert judgements rather than consensus developed through an iterative process. Second, although experts were instructed to focus on each item’s underlying concept rather than its wording or response options, the illustrative examples and response options may have influenced their ratings (i.e., introduced framing effects). Moreover, respondent burden was assessed at the item level and did not account for the cumulative burden associated with questionnaire structure and response volume. Accordingly, while our assessment considered item-level burden, overall burden may reflect the interaction between the number of contacts reported and the amount of information collected for each contact, in addition to the burden of non-contact-specific questionnaire items.

Third, the available evidence was limited for some aspects of social contact survey design. Although information on the size of the potential contact pool has been suggested as useful for understanding how contact patterns relate to population structure (27), few studies in the reviewed literature collected or analysed this variable. Consequently, its contribution to social contact survey design remains difficult to evaluate. Similarly, household-level social contact studies remain relatively uncommon. Future studies combining household-level data with information on contact location and relationship type may help improve our understanding of clustering in contact behaviour. When linked with infection data, such information may also provide a closer link between reported contacts and potential exposure, particularly in household transmission studies where contacts can be characterised in relation to an identified infected household member. Such data may also support estimation of epidemiological quantities, such as the serial interval, further motivating the need for expanded household-level data collection (89,90). Fourth, social contact surveys generally measure interactions that could potentially lead to transmission rather than contacts that have resulted in transmission. Whether a reported contact would lead to exposure or infection depends on factors that are not directly observed, including pathogen characteristics, infectiousness, and features of the interaction. Moreover, individuals may behave differently when infectious or when interacting with an infectious person, making this counterfactual relationship difficult to establish. The questionnaire proposed here partially addresses this limitation by collecting additional characteristics of contacts that may help contextualise their potential relevance to transmission. However, how these characteristics should be combined to represent variation in transmission risk across contacts remains an important area for further research. Finally, the proposed questionnaire has not yet been tested in field settings. Although the expert assessment provides an initial evaluation of perceived item importance, respondent burden, and expected reporting accuracy, the questionnaire’s practical applicability remains to be established. Future studies should assess its comprehensibility, feasibility, and respondent burden under field conditions.

## Conclusions

This study provides an expert-informed questionnaire for diary-based social contact surveys, together with updated recommendations addressing sampling, survey method, contact definitions, essential contact information, and data governance. Together, the questionnaire and recommendations aim to support more consistent, comparable, and timely social contact data collection, strengthening preparedness for future public health emergencies.

## Data Availability

All data produced in the present work are contained in the manuscript

## Authors contributions (based on CRediT Taxonomy)

Conceptualisation: NL, GL, LDD, and NH

Data curation: NL, GL, LDD, and NH

Formal analysis: NL

Methodology: NL, GL, LDD, and NH

Project administration: NL

Visualisation: NL

Supervision: NH

Writing (original draft): NL, GL, LDD, JS, PB, LW, SA and NH

Writing (review and editing): NL, GL, LDD, JS, PB, LW, SA and NH in consultation with all the other authors.

## Appendix

**Figure S1.**
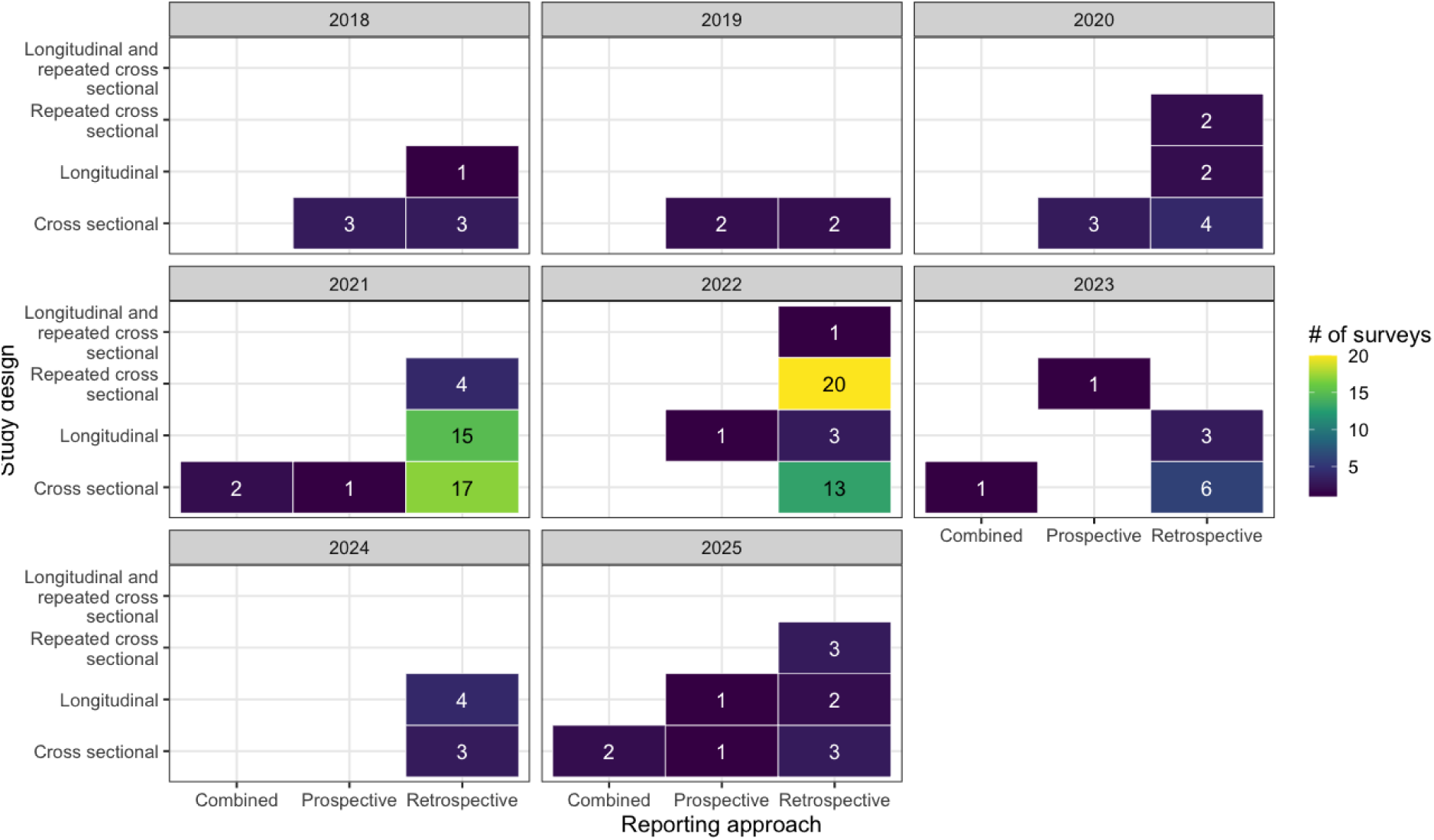
Distribution of study design and reporting approach by publication year.

**Figure S2.**
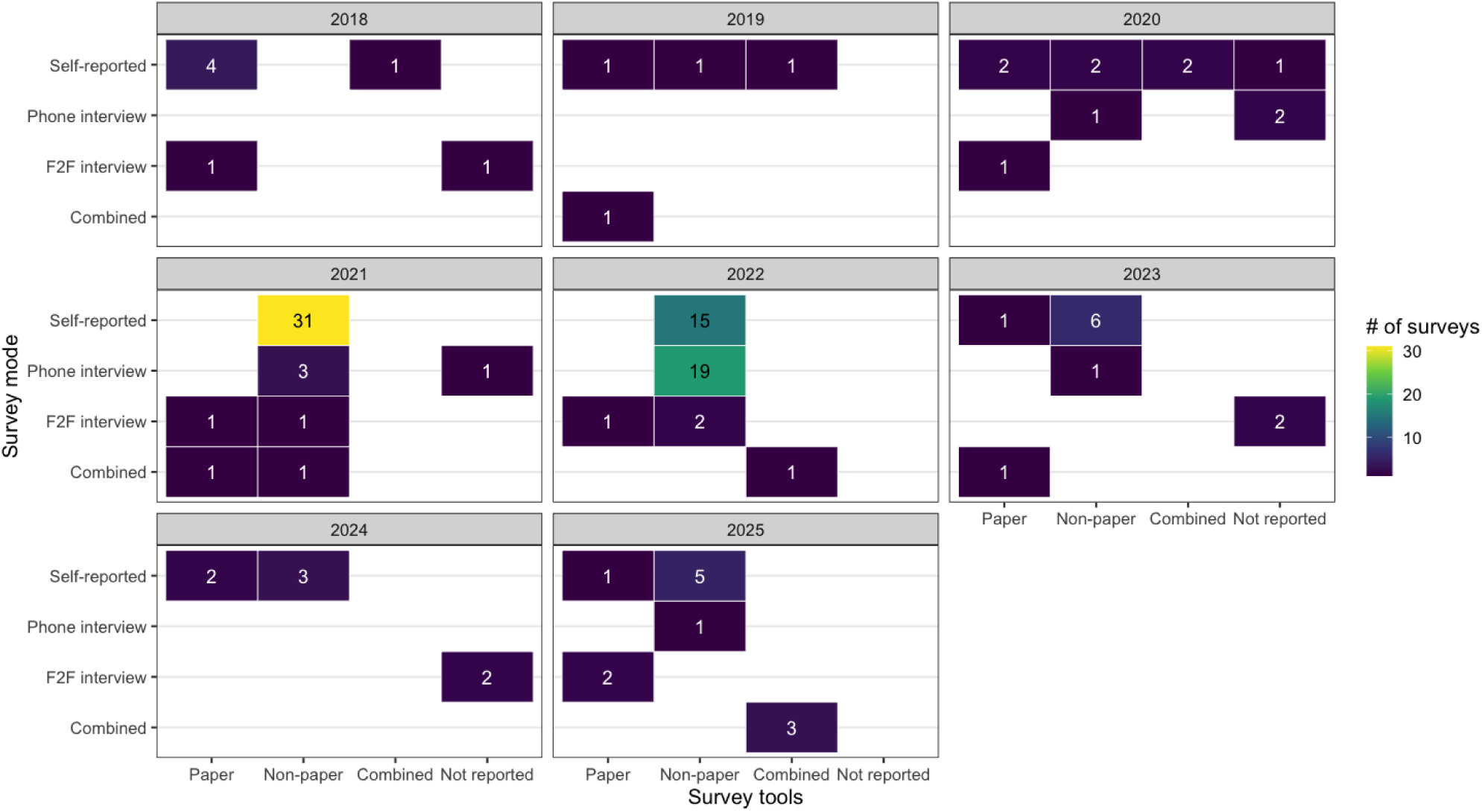
Distribution of survey mode and survey tools by publication year.

**Figure S3.**
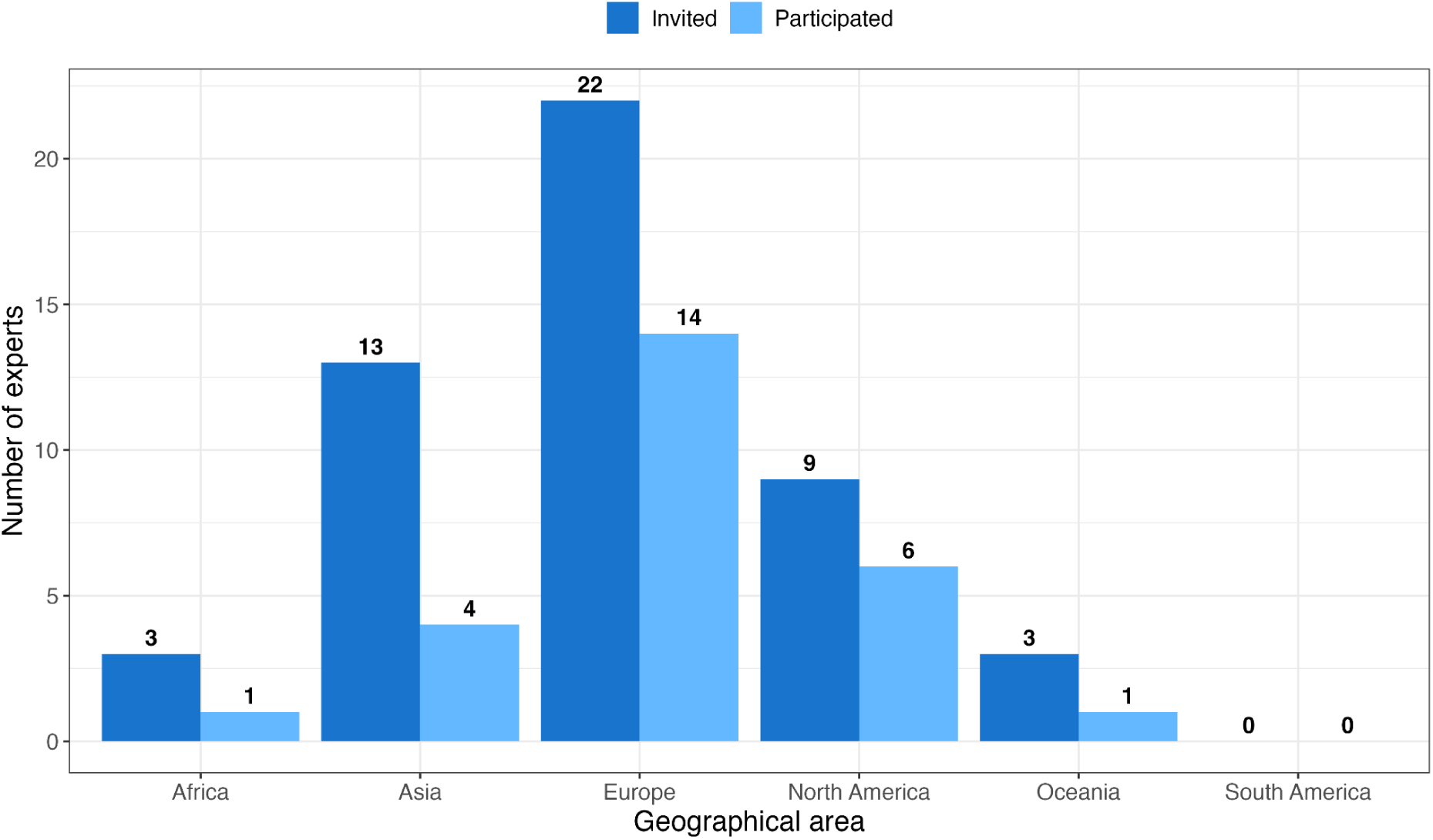
Geographic distribution of invited and participating experts by country of institutional affiliation.

**Figure S4.**
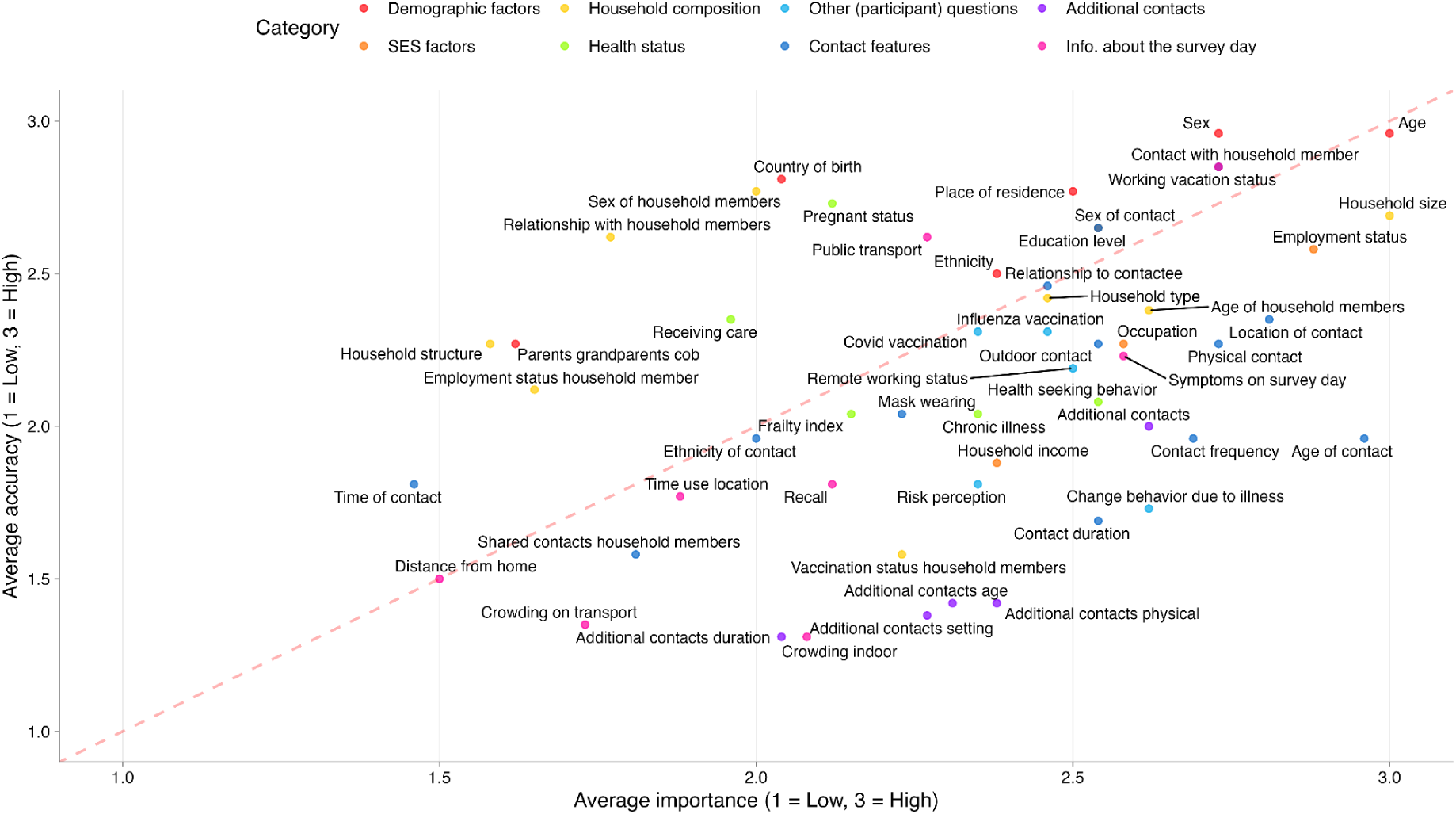
Expert assessment matrix mapped by the average importance and accuracy.

**Figure S5.**
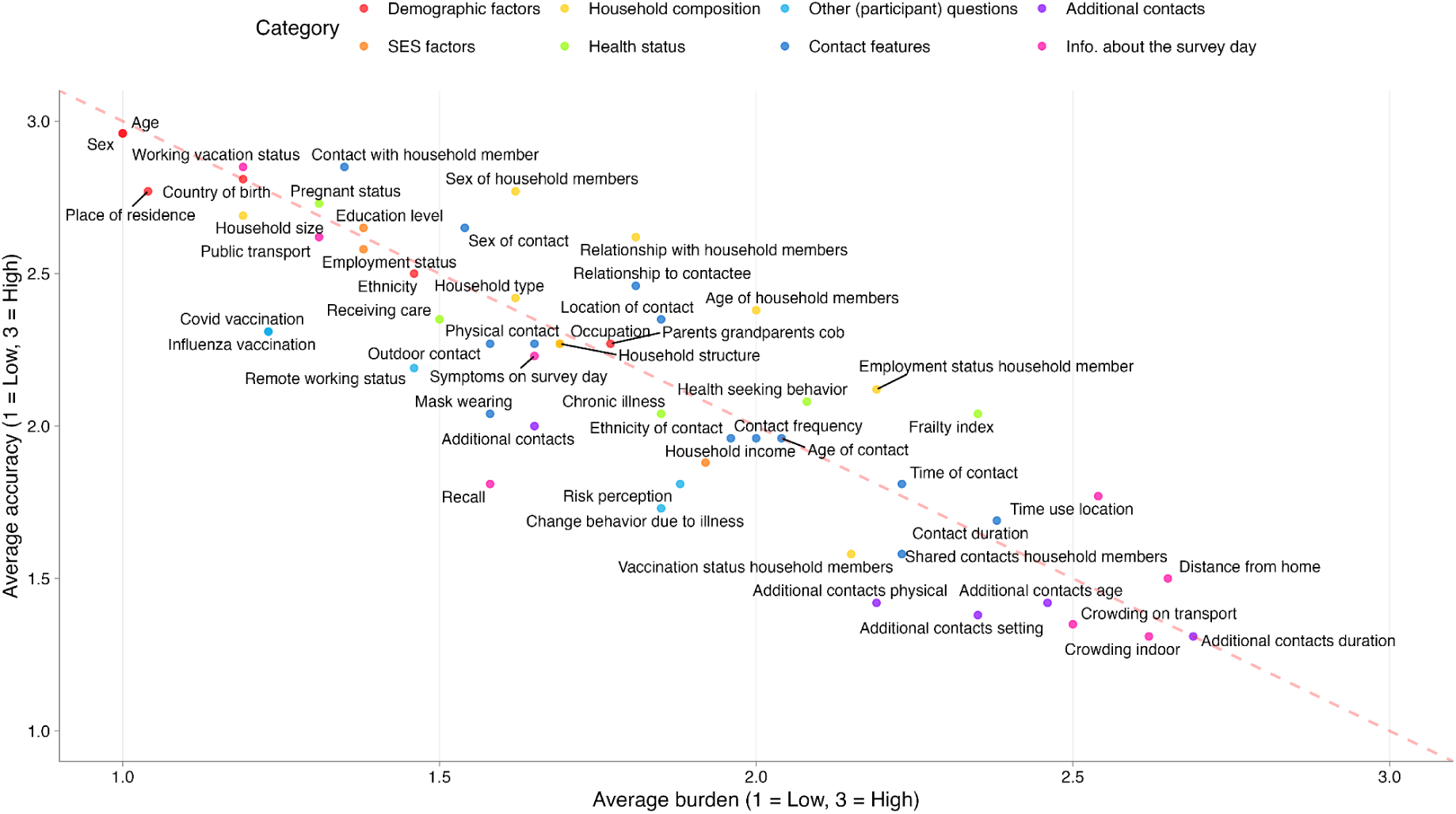
Expert assessment matrix mapped by the average reporting burden and accuracy.

**Figure S6.**
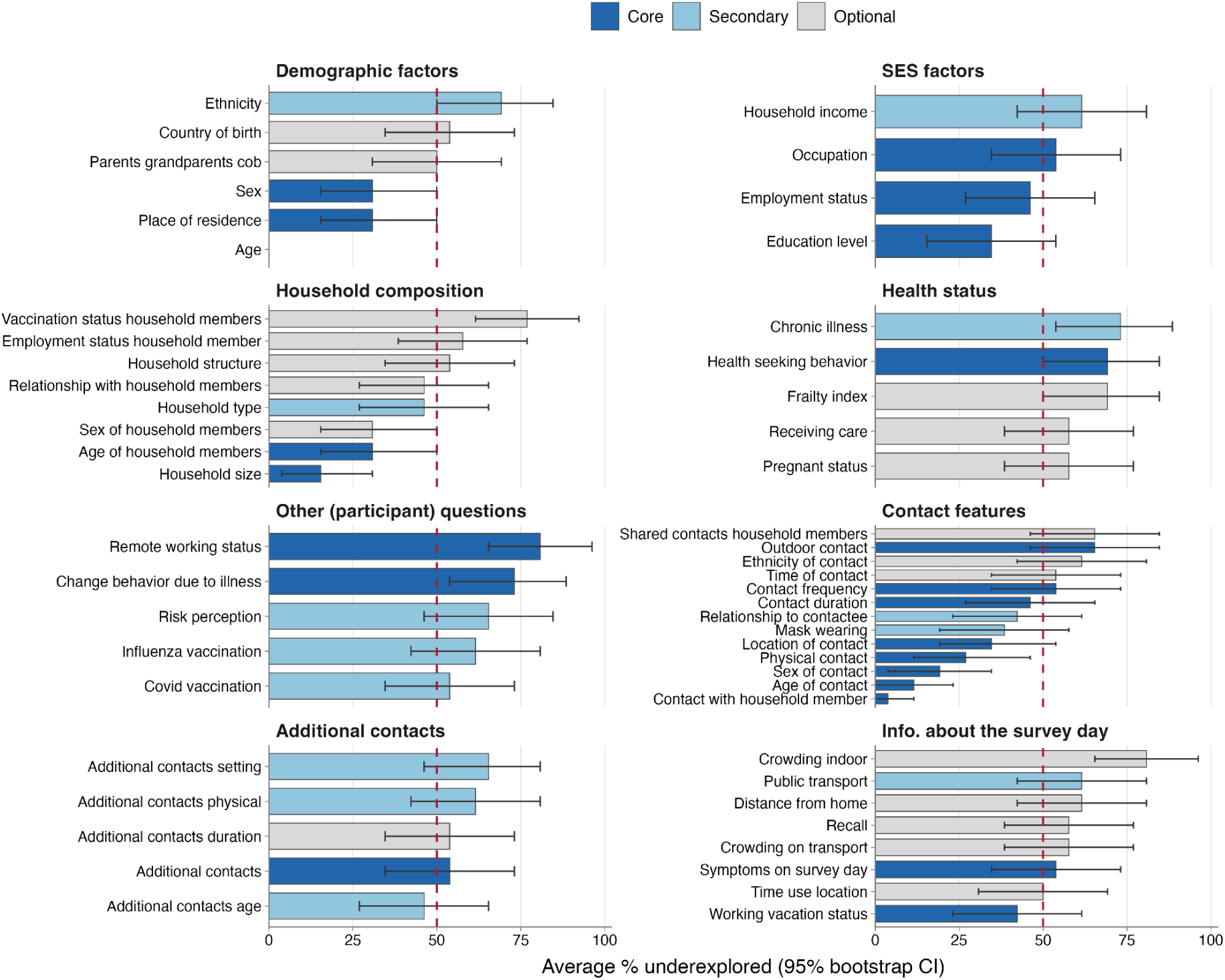
Expert-reported under exploration of survey items. Values represent the proportion of experts who indicated that further research is needed for each item.

**Figure S7.**
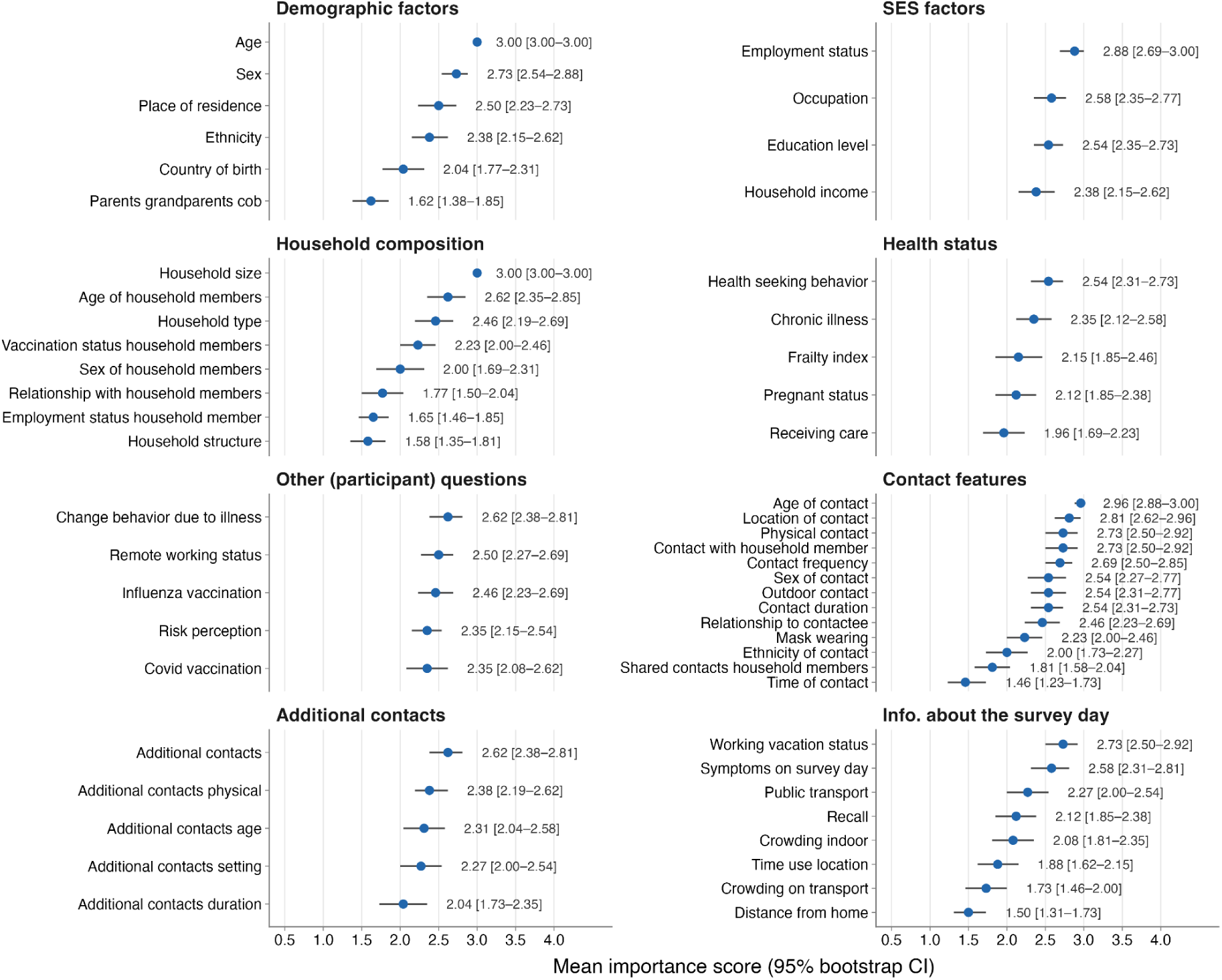
Average importance scores for each survey item, with 95% confidence intervals.

**Figure S8.**
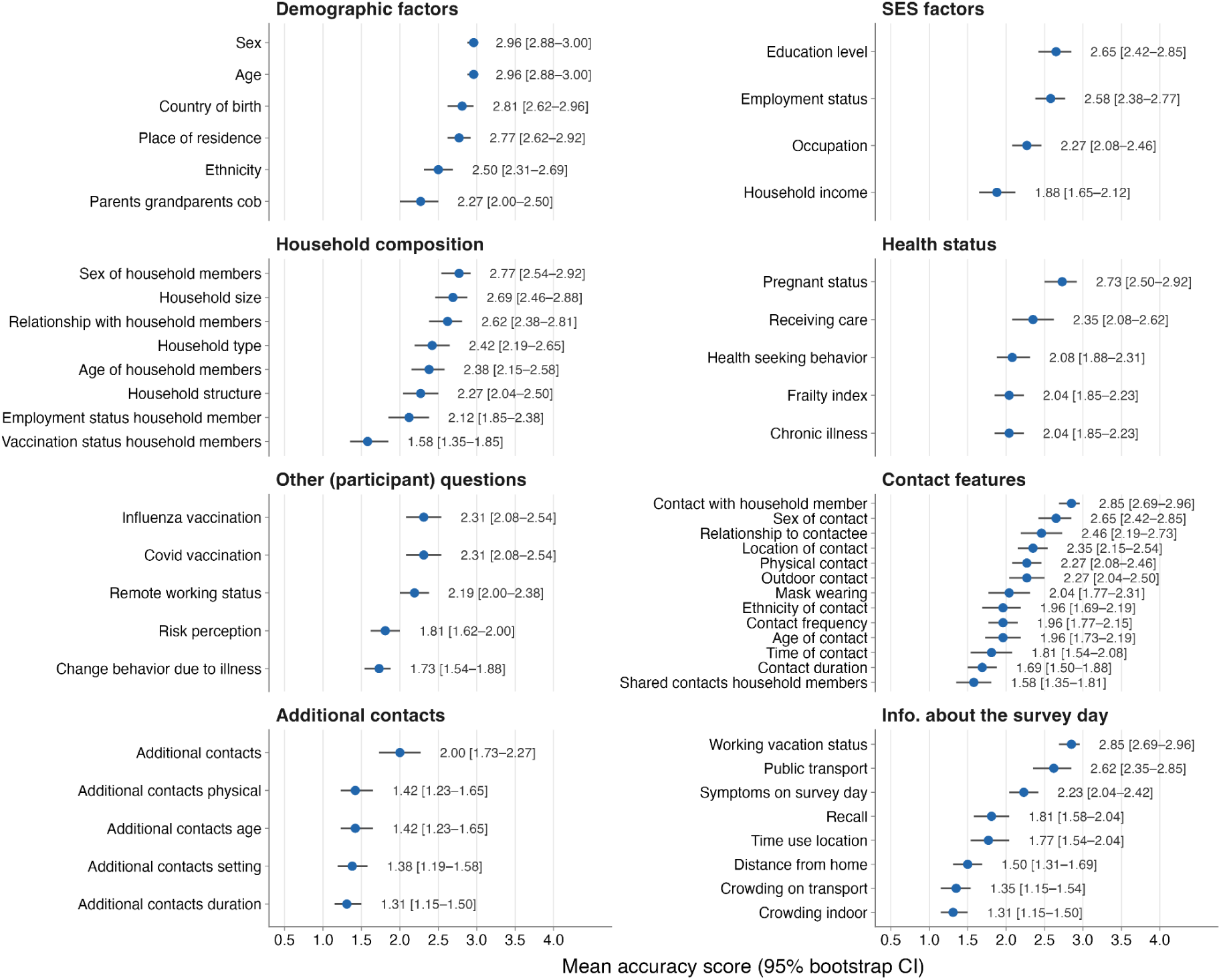
Average accuracy scores for each survey item, with 95% confidence intervals.

**Figure S9.**
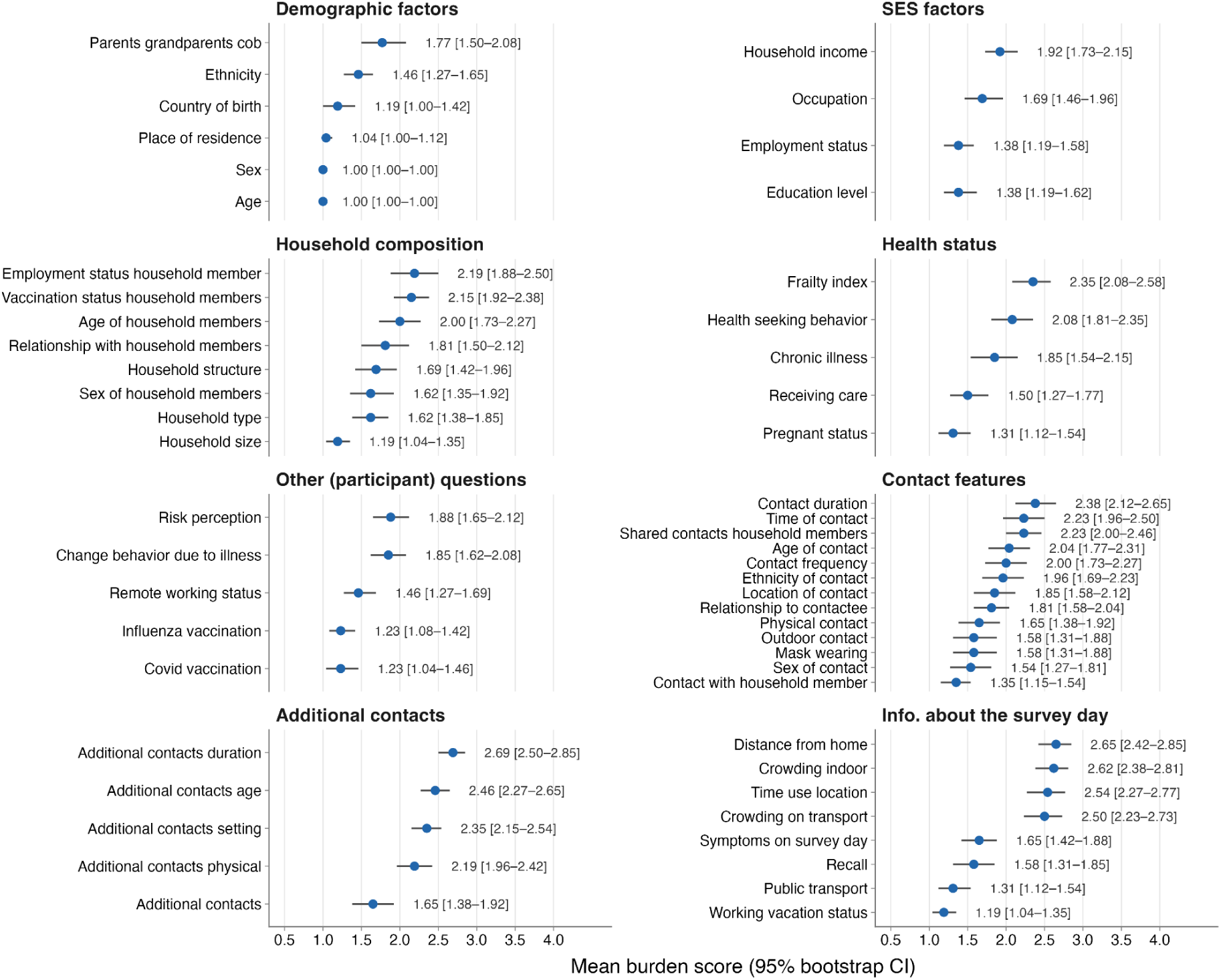
Average burden scores for each survey item, with 95% confidence intervals.

**Figure S10.**
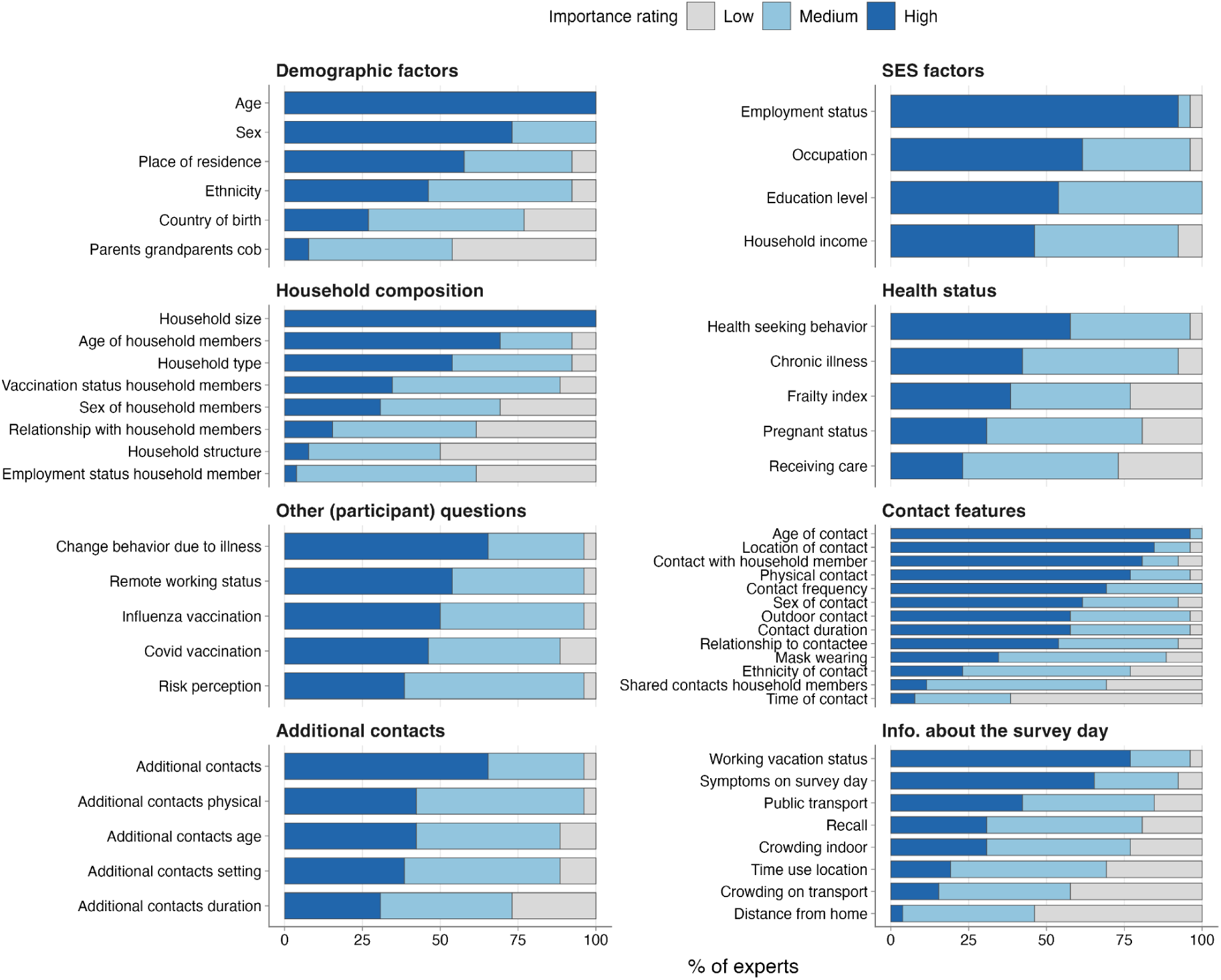
Distribution of importance ratings from experts.

**Table S1.** Database-specific search strategies used for the systematic review.

| Database | Search string |
|---|---|
| PubMed | ((panels*[Title/Abstract] OR survey*[Title/Abstract] OR questionnaire*[Title/Abstract] OR diary[Title/Abstract] OR diaries[Title/Abstract]) AND<br>(social contact*[Title/Abstract] OR mixing behavio*[Title/Abstract] OR mixing pattern*[Title/Abstract] OR contact pattern*[Title/Abstract] OR contact network*[Title/Abstract] OR contact survey*[Title/Abstract] OR contact data[Title/Abstract] OR contact mixing [Title/Abstract] OR contact rate*[Title/Abstract] OR contact matri*[Title/Abstract] OR contact parameter*[Title/Abstract] OR physical contact*[Title/Abstract] OR social mixing[Title/Abstract] OR mixing matri*[Title/Abstract] OR assortative mixing[Title/Abstract] OR disassortative mixing[Title/Abstract] OR mixing parameter*[Title/Abstract])) |
| Web of Science | (TI=(panels* OR survey* OR questionnaire* OR diary OR diaries)<br>OR<br>AB=(panels* OR survey* OR questionnaire* OR diary OR diaries)) |
|  | AND<br>(TI=(social contact* OR mixing behavio* OR mixing pattern* OR contact pattern* OR contact network* OR contact survey* OR contact data OR contact mixing OR contact rate* OR contact matri* OR contact parameter* OR physical contact* OR social mixing OR mixing matri* OR assortative mixing OR disassortative mixing OR mixing parameter*)<br>OR<br>AB=(social contact* OR mixing behavio* OR mixing pattern* OR contact pattern* OR contact network* OR contact survey* OR contact data OR contact mixing OR contact rate* OR contact matri* OR contact parameter* OR physical contact* OR social mixing OR mixing matri* OR assortative mixing OR disassortative mixing OR mixing parameter*)) |

**Table S2.** Survey-level distribution of study characteristics and initiative-level sensitivity analysis. Initiative-level counts use 83 unique initiatives, with surveys from the same initiative merged (i.e., Del Fava et al. (2021), Dobreva et al. (2022), and CoMix studies (16,34,37,38,91–96)). Each initiative is counted once in every distinct category represented among its constituent surveys. Categories may overlap for geographical area and study design; therefore, percentages may exceed 100%.

| Characteristics | Category | Survey-level entries: n/N (%) | Initiative-level entries: n/N (%) |
|---|---|---|---|
| Geographical area | Europe | 51/129 (39.5%) | 24/83 (28.9%) |
|  | Asia | 29/129 (22.5%) | 29/83 (34.9%) |
|  | Africa | 28/129 (21.7%) | 10/83 (12.0%) |
|  | North America | 18/129 (13.9%) | 18/83 (21.7%) |
|  | Oceania | 2/129 (1.5%) | 2/83 (2.4%) |
|  | South America | 1/129 (0.7%) | 1/83 (1.2%) |
| Study design | Cross-sectional | 66/129 (51.2%) | 58/83 (69.9%) |
|  | Longitudinal | 32/129 (24.8%) | 13/83 (15.7%) |
|  | Repeated cross-sectional | 30/129 (23.3%) | 13/83 (15.7%) |
|  | Combined longitudinal and repeated cross-sectional | 1/129 (0.8%) | 1/83 (1.2%) |
| Reporting approach | Retrospective | 111/129 (86.0%) | 65/83 (78.3%) |
|  | Prospective | 13/129 (10.1%) | 13/83 (15.7%) |
|  | Combined retrospective and prospective | 5/129 (3.9%) | 5/83 (6.1%) |
| Survey mode | Self-administered | 79/129 (61.2%) | 51/83 (61.4%) |
|  | Phone interview | 28/129 (21.7%) | 10/83 (12.0%) |
|  | Face-to-face interview | 14/129 (10.9%) | 14/83 (16.9%) |
|  | Mixed modes | 8/129 (6.2%) | 8/83 (9.6%) |
| Sampling design | Non-probability | 100/129 (77.5%) | 56/83 (67.5%) |
|  | Probability | 28/129 (21.7%) | 26/83 (31.3%) |
|  | Combined probability and non-probability | 1/129 (0.8%) | 1/83 (1.2%) |
| Sampling stages | One-stage | 101/129 (78.3%) | 55/83 (66.3%) |
|  | Multistage | 27/129 (20.9%) | 27/83 (32.5%) |
|  | Combined one-stage and multistage | 1/129 (0.8%) | 1/83 (1.2%) |
| Contact reporting level | Individual-level | 120/129 (93.0%) | 74/83 (89.2%) |
|  | Household-level | 9/129 (7.0%) | 9/83 (10.8%) |
| Contact definition | Physical or conversational | 108/129 (83.7%) | 62/83 (74.7%) |
|  | Proximity-based | 7/129 (5.4%) | 7/83 (8.4%) |
|  | Physical, conversational, and proximity-based | 7/129 (5.4%) | 7/83 (8.4%) |
|  | Conversational only | 4/129 (3.1%) | 4/83 (4.8%) |
|  | Physical only | 2/129 (1.6%) | 2/83 (2.4%) |
|  | Physical or proximity-based | 1/129 (0.8%) | 1/83 (1.2%) |

**Table S3.**
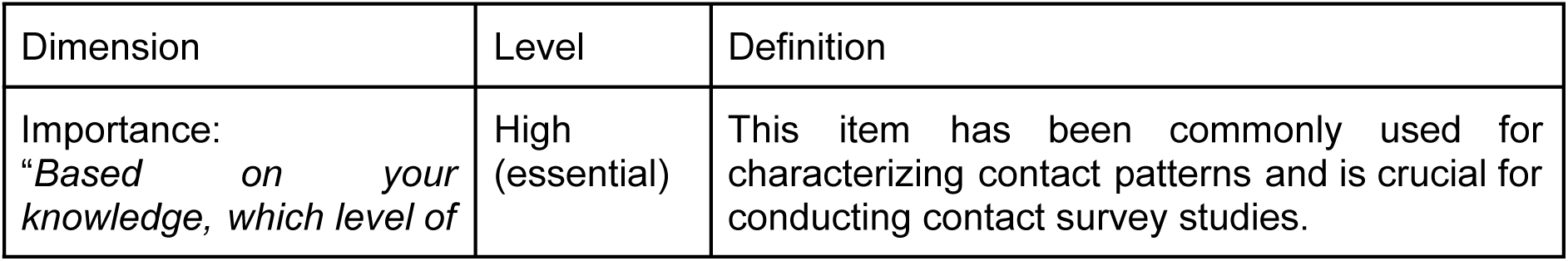
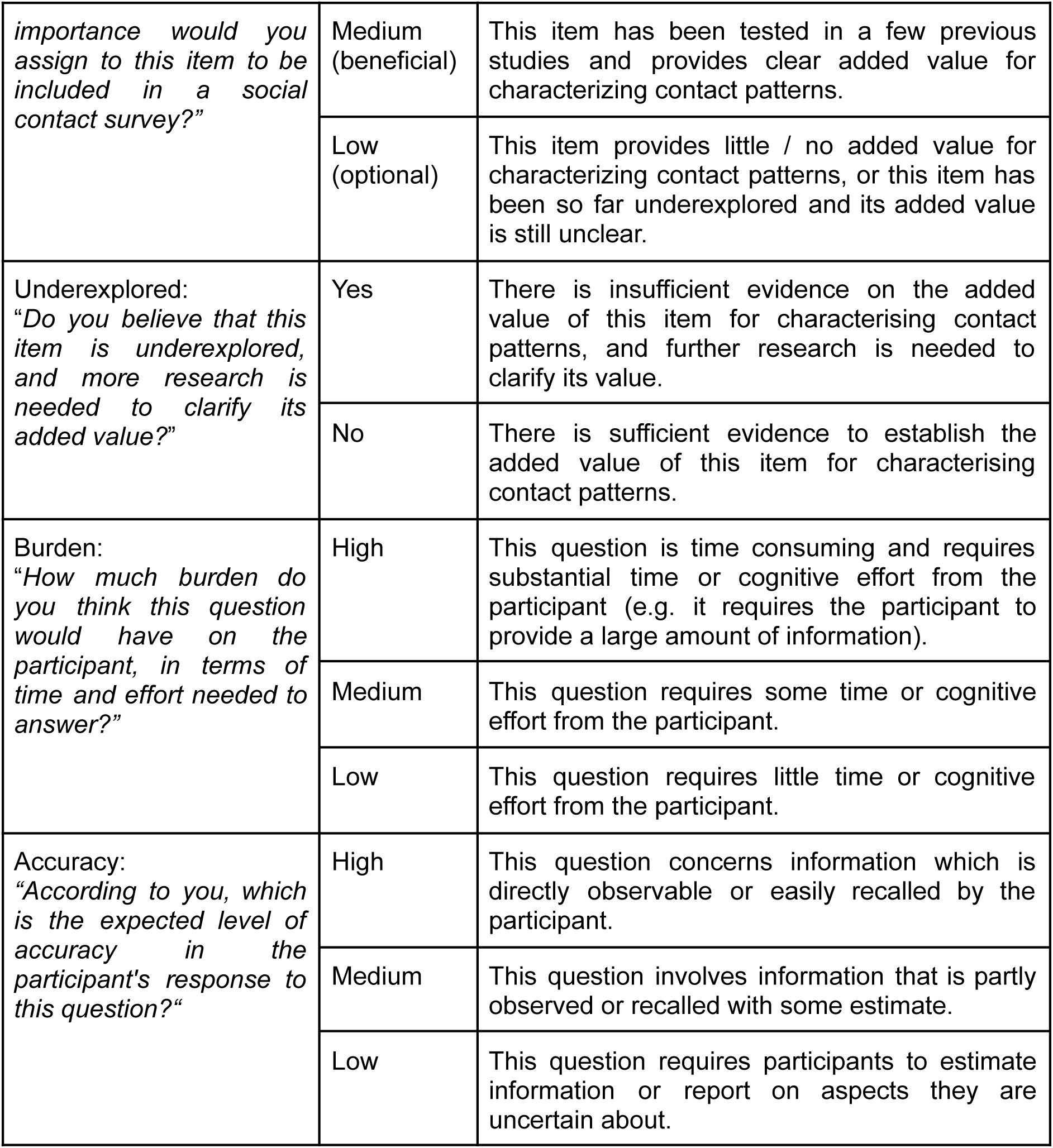
Definitions of expert assessment dimensions and response categories.

**Table S4.** Average (together with 95% bootstrap-based CIs) expert ratings of importance, accuracy, burden, and underexplored across survey thematic categories.

| Category | Importance | Accuracy | Burden | Underexplored (%) |
|---|---|---|---|---|
| Demographic factors (n = 6) | 2.38 (2.26–2.49) | **2.71** (2.63–2.79) | 1.24 (1.17–1.33) | 39.1 (31.4–46.8) |
| SES factors (n = 4) | **2.60** (2.48–2.70) | 2.35 (2.23–2.46) | 1.60 (1.48–1.71) | 49 (39.4–58.7) |
| Household composition (n = 8) | 2.16 (2.06–2.26) | 2.36 (2.26–2.45) | 1.78 (1.68–1.88) | 44.7 (38–51.4) |
| Health status (n = 5) | 2.22 (2.10–2.35) | 2.25 (2.15–2.35) | 1.82 (1.68–1.95) | 65.4 (56.9–73.8) |
| Other (participant) questions (n = 5) | 2.45 (2.35–2.55) | 2.07 (1.97–2.17) | 1.53 (1.42–1.65) | **66.9 (58.5–74.6)** |
| Contact features (n = 13) | 2.42 (2.35–2.50) | 2.14 (2.07–2.22) | 1.86 (1.78–1.94) | 40.2 (34.9–45.6) |
| Additional contacts (n = 5) | 2.32 (2.21–2.44) | 1.51 (1.40–1.62) | **2.27** (2.15–2.38) | 56.2 (47.7–64.6) |
| Information about the survey day (n = 8) | 2.11 (2.01–2.22) | 1.93 (1.83–2.03) | 2.00 (1.89–2.12) | 58.2 (51.4–64.9) |

