## Supplementary Material 5 for "On conducting diary-based social contact surveys to inform transmission models of close-contact infections: a systematic review and expert-informed questionnaire"

### Social contact survey items

**AIM & MOTIVATION**

We aim to develop **a standardised protocol for conducting a survey on social contacts** to inform transmission models of close-contact infections.

We conducted a systematic review of studies collecting primary data on social contacts between humans, published in 2018-2025. Based on the questionnaires available, we extracted a comprehensive list of possible survey questions used in studies conducted in the general population (i.e. not in confined settings). The objective of this survey is to **gather expert opinion** on which questions are essential for inclusion in a contact questionnaire and which questions may provide additional value for improving the characterisation of mixing patterns and disease spread.

**STUDY INFORMATION AND CONSENT**

This survey is intended for researchers with experience in social contact data collection, infectious disease epidemiology, or related fields. Participation consists in evaluating each survey item based on a set of pre-defined metrics, and is expected to take approximately **20–30 minutes**.

Responses will be analysed in an aggregated form for research purposes and may be reported in scientific publications or presentations. Survey responses will be treated confidentially and will be accessible only to the research team. Participation is voluntary, and you may stop at any time before submitting your responses.

The full informed consent form is made available to participants via this link.

* Indicates required question

1. Email *

______________________________

2. Informed consent *

*Tick all that apply.*

☐ By proceeding with this survey, I declare that I have been informed of and understand the purpose of the study, its duration and consequences, any risks and inconveniences, and what is expected of me. I agree to participate in the study as is described in the informed consent form.

#### Evaluation framework

For each item, we kindly ask you to provide an evaluation using three metrics: **Importance**, **Burden**, and **Accuracy**, as defined below. When providing your evaluation, please focus on the **topic of the question**, rather than on the specific wording or the answer options. These definitions will be displayed at the top of each survey section to guide the evaluation.

**1. Importance:** *Based on your knowledge, which level of importance would you assign to this item to be included in a social contact survey?*

- **HIGH (Essential):** *this item is crucial for contact survey studies, and has been commonly used for characterizing contact patterns.*
- **MEDIUM (Beneficial):** *this item has been tested in a few previous studies and provides clear added value for characterizing contact patterns.*
- **LOW (Optional):** *this item provides little / no added value for characterizing contact patterns, or this item has been so far underexplored and its added value is still unclear.*

***Additionally:*** *Do you believe that this item is underexplored, and more research is needed to clarify their added value?* ***YES/NO***

**2. Burden:** *How much burden do you think this question would have on the participant, in terms of time and effort needed to answer?*

- **HIGH**: *this question is time consuming and requires a lot of effort / a lot of information from the participant.*
- **MEDIUM**: *this question requires some time / some effort from the participant.*
- **LOW**: *this question does not require a lot of effort / time from the participant.*

**3. Accuracy:** *According to you, which is the expected level of accuracy in the participant's response to this question?*

- **HIGH**: *this question concerns information which is directly observable or easily recalled by the participant.*
- **MEDIUM**: *this question involves information that is partly observed or recalled with some estimate.*
- **LOW**: *this question requires participants to estimate information or report on aspects they are uncertain about.*

#### Instructions

When evaluating each survey item, please use the guidance below.

**1. Survey context**

Assume an adult participant from the general population taking part in a cross-sectional, retrospective study. Data are collected using a paper-based contact diary, in which participants report their own contacts from the previous day.

**2. Geographical context**

Base the evaluation on the country in which you currently live. Some answer options may reflect specific country contexts and are provided for illustrative purposes only; items should be evaluated as if adapted to the country-specific context.

**3. Disease context**

The questionnaire is meant to be as general as possible, and not targeted to a specific disease. Provide your evaluation based on the disease(s) most frequently studied in your research. If the perceived importance of an item varies by disease, adopt a conservative approach (e.g., rate as "high importance" if the metric's criteria are met for at least one disease).

**4. Burden assessment**

Burden should be evaluated per individual survey item and is not cumulative across the questionnaire. Assess the burden of each item independently, regardless of the burden imposed by preceding or subsequent items. When evaluating burden, focus on the topic of the question rather than on the specific answer options, which are illustrative.

**5. Repeated and combined items**

Some survey items may be asked multiple times (e.g. once per household member or per contact). In such cases, provide the evaluation as if the item were asked only once, without considering the cumulative burden of repeated administration. Items consisting of multiple subquestions should be evaluated as a single, combined item.

The instructions are also available here, so you can access them any time while you are filling the survey.

#### SECTION 1: participant's background questions

This section includes questions on participant background, including demographic and socio-economic characteristics, household composition, and health status.

#### Demographic factors

If you want to review the instructions or the definition of the evaluation metrics, you can find them here.

3. **Q1: age** *
What is your age?
[insert number]

*Mark only one oval per row.*

| | Low | Medium | High |
|---|---|---|---|
| **Importance** | | | |
| **Burden** | | | |
| **Accuracy** | | | |

4. **Q1: age** *
Do you believe that this item is underexplored, and more research is needed to clarify their added value?

*Mark only one oval.*

- Yes
- No

5. **Q2: sex** *
What is your sex?
☐ female
☐ male
☐ prefer not to answer

*Mark only one oval per row.*

| | Low | Medium | High |
|---|---|---|---|
| **Importance** | | | |
| **Burden** | | | |
| **Accuracy** | | | |

6. **Q2: sex** *
Do you believe that this item is underexplored, and more research is needed to clarify their added value?

*Mark only one oval.*

- Yes
- No

7. **Q3: place of residence** *
What is the postal code of your place of residence?
[insert number]

*Mark only one oval per row.*

| | Low | Medium | High |
|---|---|---|---|
| **Importance** | | | |
| **Burden** | | | |
| **Accuracy** | | | |

8. **Q3: place of residence** *
Do you believe that this item is underexplored, and more research is needed to clarify their added value?

*Mark only one oval.*

- Yes
- No

9. **Q4: country of birth** *
In which country were you born?
☐ In Belgium [or country of study]
☐ In another European country
☐ In a country outside Europe

*Mark only one oval per row.*

| | Low | Medium | High |
|---|---|---|---|
| **Importance** | | | |
| **Burden** | | | |
| **Accuracy** | | | |

10. **Q4: country of birth** *
Do you believe that this item is underexplored, and more research is needed to clarify their added value?

*Mark only one oval.*

- Yes
- No

11. **Q5: ethnicity** *

Which ethnic group do you belong to?

- White
- ☐ English / Welsh / Scottish / Northern Irish / British
- ☐ Irish
- ☐ Gypsy or Irish Traveller
- ☐ Any other White background

- Mixed / multiple ethnic groups
- ☐ White and Black Caribbean
- ☐ White and Black African
- ☐ White and Asian
- ☐ Any other Mixed / multiple ethnic background

- Asian / Asian British
- ☐ Indian
- ☐ Pakistani
- ☐ Bangladeshi
- ☐ Chinese
- ☐ Any other Asian background

- Black / African / Caribbean / Black British
- ☐ African
- ☐ Caribbean
- ☐ Any other Black / African / Caribbean background

- Other ethnic group
- ☐ Arab
- ☐ Any other ethnic group, please write in ___________
- ☐ Prefer not to answer

*Mark only one oval per row.*

| | Low | Medium | High |
|---|---|---|---|
| **Importance** | ◯ | ◯ | ◯ |
| **Burden** | ◯ | ◯ | ◯ |
| **Accuracy** | ◯ | ◯ | ◯ |

12. **Q5: ethnicity** *

Do you believe that this item is underexplored, and more research is needed to clarify their added value?

*Mark only one oval.*

◯ Yes

◯ No

13. **Q6: parents' & grandparents' country of birth** *

In which country was your mother born?
- [ ] In Belgium [or country of study]
- [ ] In another European country
- [ ] In a country outside Europe

In which country was your father born?
- [ ] In Belgium [or country of study]
- [ ] In another European country
- [ ] In a country outside Europe

In which country was you grandfather/grandmother from father/mother side born? [4 subquestions]
- [ ] In Belgium [or country of study]
- [ ] In another European country
- [ ] In a country outside Europe

*Mark only one oval per row.*

| | Low | Medium | High |
|---|---|---|---|
| **Importance** | ◯ | ◯ | ◯ |
| **Burden** | ◯ | ◯ | ◯ |
| **Accuracy** | ◯ | ◯ | ◯ |

14. **Q6: parents' & grandparents' country of birth** *

Do you believe that this item is underexplored, and more research is needed to clarify their added value?

*Mark only one oval.*

◯ Yes

◯ No

#### Socio-economic factors

If you want to review the instructions or the definition of the evaluation metrics, you can find them here.

15. **Q7: education level** *

What is the highest level of education you have completed?

- ☐ Without a diploma or primary education
- ☐ General lower secondary education (first 3 years completed)
- ☐ Technical, artistic or professional lower secondary education (first 3 years completed)
- ☐ General upper secondary education (6 years completed)
- ☐ Technical or artistic upper secondary education (6 years)
- ☐ Professional upper secondary (6 years)
- ☐ Higher education: graduat, candidature, bachelor
- ☐ University education: bachelor's degree, post-graduate, master's degree
- ☐ Complementary master
- ☐ Doctorate

*Mark only one oval per row.*

| | Low | Medium | High |
|---|---|---|---|
| **Importance** | ⬭ | ⬭ | ⬭ |
| **Burden** | ⬭ | ⬭ | ⬭ |
| **Accuracy** | ⬭ | ⬭ | ⬭ |

16. **Q7: education level** *

Do you believe that this item is underexplored, and more research is needed to clarify their added value?

*Mark only one oval.*

⬭ Yes

⬭ No

17. **Q8: household income** *

Into which category does your total net household monthly income from all sources fall (i.e. income after tax deduction)?

*Please include all your income sources: salaries, scholarships, pension and Social Security benefits, dividends from shares, income from rental properties, child support and alimony etc.*

- ☐ € 0 - € 549
- ☐ € 550 - € 999
- ☐ € 1 000 - € 1 299
- ☐ € 1 300 - € 1 499

... ... ...

- ☐ € 4 500 - € 5 499
- ☐ € 5 500 - € 7 999
- ☐ € 8 000 or more
- ☐ Prefer not to answer

*Mark only one oval per row.*

| | Low | Medium | High |
|---|---|---|---|
| **Importance** | ◯ | ◯ | ◯ |
| **Burden** | ◯ | ◯ | ◯ |
| **Accuracy** | ◯ | ◯ | ◯ |

18. **Q8: household income** *

Do you believe that this item is underexplored, and more research is needed to clarify their added value?

*Mark only one oval.*

- ◯ Yes
- ◯ No

19. **Q9: employment status** *

What is your current employment status?

- ☐ Employed full-time (34 hours or more)
- ☐ Employed part-time (less than 34 hours)
- ☐ Self employed
- ☐ Unemployed but looking for a job
- ☐ Unemployed and not looking for a job
- ☐ Full-time parent, homemaker
- ☐ Retired
- ☐ Student/Pupil
- ☐ Long-term sick or disabled

*Mark only one oval per row.*

| | Low | Medium | High |
|---|---|---|---|
| **Importance** | ◯ | ◯ | ◯ |
| **Burden** | ◯ | ◯ | ◯ |
| **Accuracy** | ◯ | ◯ | ◯ |

20. **Q9: employment status** *

Do you believe that this item is underexplored, and more research is needed to clarify their added value?

*Mark only one oval.*

◯ Yes

◯ No

21. **Q10: occupation** *

What is your occupation?

Self-employed

- ☐ Craftperson, trader with no employees
- ☐ Craftperson, trader with 5 employees or less
- ☐ Business owner, trader with 6 employees or more
- ☐ Liberal profession

Employee

- ☐ Member of the executive management, senior management
- ☐ Middle management, not part of executive management
- ☐ Other employees

Manual worker

- ☐ Worker with vocational training
- ☐ Worker without vocational training

Other

- ☐ Homemaker
- ☐ Person with a disability
- ☐ Retired
- ☐ Student
- ☐ Unemployed
- ☐ Rentier

*Mark only one oval per row.*

| | Low | Medium | High |
|---|---|---|---|
| **Importance** | ◯ | ◯ | ◯ |
| **Burden** | ◯ | ◯ | ◯ |
| **Accuracy** | ◯ | ◯ | ◯ |

22. **Q10: occupation** *

Do you believe that this item is underexplored, and more research is needed to clarify their added value?

*Mark only one oval.*

◯ Yes

◯ No

#### Household composition

If you want to review the instructions or the definition of the evaluation metrics, you can find them here.

23. **Q11: household size** *

Including you, how many other people live in your household?

*By household, we mean anyone living at the same address as you, that you share a kitchen with.*
[insert number]

*Mark only one oval per row.*

| | Low | Medium | High |
|---|---|---|---|
| **Importance** | ⬭ | ⬭ | ⬭ |
| **Burden** | ⬭ | ⬭ | ⬭ |
| **Accuracy** | ⬭ | ⬭ | ⬭ |

24. **Q11: household size** *

Do you believe that this item is underexplored, and more research is needed to clarify their added value?

*Mark only one oval.*

- Yes
- No

25. **Q12: household type** *

Which of the following best describes your household?

House or apartment
- ☐ Two or more non-family adults
- ☐ Couple with dependent children
- ☐ Couple with independent children only
- ☐ Couple with no children
- ☐ Lone parent with dependent children
- ☐ Lone parent with independent children only
- ☐ Households containing two or more families

Collective setting
- ☐ Hotel/hostel
- ☐ Student residence
- ☐ Nursing home
- ☐ Other

*Mark only one oval per row.*

| | Low | Medium | High |
|---|---|---|---|
| **Importance** | ⬭ | ⬭ | ⬭ |
| **Burden** | ⬭ | ⬭ | ⬭ |
| **Accuracy** | ⬭ | ⬭ | ⬭ |

26. **Q12: household type** *
Do you believe that this item is underexplored, and more research is needed to clarify their added value?

*Mark only one oval.*

- Yes
- No

27. **Q13: age of household members** *
[ASK for each household member]

What is their age?
[insert number/age group]

*Mark only one oval per row.*

| | Low | Medium | High |
|---|---|---|---|
| **Importance** | | | |
| **Burden** | | | |
| **Accuracy** | | | |

28. **Q13: age of household members** *
Do you believe that this item is underexplored, and more research is needed to clarify their added value?

*Mark only one oval.*

- Yes
- No

29. **Q14: sex of household members** *
[ASK for each household member]

What is their sex?
☐ female
☐ male
☐ prefer not to answer

*Mark only one oval per row.*

| | Low | Medium | High |
|---|---|---|---|
| **Importance** | | | |
| **Burden** | | | |
| **Accuracy** | | | |

30. **Q14: sex of household members** *

Do you believe that this item is underexplored, and more research is needed to clarify their added value?

*Mark only one oval.*

- Yes
- No

31. **Q15: relationship with household members** *

[ASK for each household member]

How is this member related to you?
☐ Spouse
☐ Sibling
☐ Child
☐ Grandparent
☐ Uncle/Aunt
☐ Grandchild
☐ Niece/Nephew
☐ Other Relative
☐ Not Related

*Mark only one oval per row.*

| | Low | Medium | High |
|---|---|---|---|
| **Importance** | | | |
| **Burden** | | | |
| **Accuracy** | | | |

32. **Q15: relationship with household members** *

Do you believe that this item is underexplored, and more research is needed to clarify their added value?

*Mark only one oval.*

- Yes
- No

33. **Q16: employment status of household members** *
[ASK for each household member]

What is their current employment status?
☐ Employed full-time (34 hours or more)
☐ Employed part-time (less than 34 hours)
☐ Self employed
☐ Unemployed but looking for a job
☐ Unemployed and not looking for a job
☐ Full-time parent, homemaker
☐ Retired
☐ Student/Pupil
☐ Long-term sick or disabled

*Mark only one oval per row.*

| | Low | Medium | High |
|---|---|---|---|
| **Importance** | ◯ | ◯ | ◯ |
| **Burden** | ◯ | ◯ | ◯ |
| **Accuracy** | ◯ | ◯ | ◯ |

34. **Q16: employment status of household members** *
Do you believe that this item is underexplored, and more research is needed to clarify their added value?

*Mark only one oval.*

◯ Yes

◯ No

35. **Q17: vaccination status of household members** *
[ASK for each household member]

Did they receive a vaccine in the last year/in the last winter campaign?
☐ No
☐ Yes, for COVID-19
☐ Yes, for influenza
☐ Yes, other

*Mark only one oval per row.*

| | Low | Medium | High |
|---|---|---|---|
| **Importance** | ◯ | ◯ | ◯ |
| **Burden** | ◯ | ◯ | ◯ |
| **Accuracy** | ◯ | ◯ | ◯ |

36. **Q17: vaccination status of household members** *
Do you believe that this item is underexplored, and more research is needed to clarify their added value?

*Mark only one oval.*

- Yes
- No

37. **Q18: house structure** *
What is the total number of rooms in your house?
[insert number]
How many rooms are used to sleep in your house?
[insert number]

*Mark only one oval per row.*

| | Low | Medium | High |
|---|---|---|---|
| **Importance** | | | |
| **Burden** | | | |
| **Accuracy** | | | |

38. **Q18: house structure** *
Do you believe that this item is underexplored, and more research is needed to clarify their added value?

*Mark only one oval.*

- Yes
- No

#### Health status

If you want to review the instructions or the definition of the evaluation metrics, you can find them here.

39. **Q19: health seeking behavior** *

Did you have any of the following symptoms in the last 7 days?

- [ ] Fever or high temperature
- [ ] A cough that has lasted for at least several hours
- [ ] Shortness of breath
- [ ] Aches and pains, e.g. in back, neck, shoulders or joints
- [ ] Blocked nose
- [ ] Sore throat
- [ ] Feeling unusually tired
- [ ] None of these
- [ ] Don't know
- [ ] Prefer not to answer

Have you done any of the following for these symptoms?

- [ ] Phoned medical emergency number
- [ ] Phoned a GP practice/GP out of hours service
- [ ] Visited a GP practice/GP out of hours service
- [ ] Visited Accident & Emergency (A&E)
- [ ] Visited a testing location somewhere different to these services
- [ ] Been admitted to hospital
- [ ] Don't know
- [ ] None of these
- [ ] Prefer not to answer

*Mark only one oval per row.*

| | Low | Medium | High |
|---|---|---|---|
| **Importance** | ○ | ○ | ○ |
| **Burden** | ○ | ○ | ○ |
| **Accuracy** | ○ | ○ | ○ |

40. **Q19: health seeking behavior** *

Do you believe that this item is underexplored, and more research is needed to clarify their added value?

*Mark only one oval.*

○ Yes

○ No

41. **Q20: pregnant status** *

Are you currently pregnant?

- [ ] Yes
- [ ] No
- [ ] Prefer not to answer

*Mark only one oval per row.*

| | Low | Medium | High |
|---|---|---|---|
| **Importance** | | | |
| **Burden** | | | |
| **Accuracy** | | | |

42. **Q20: pregnant status** *

Do you believe that this item is underexplored, and more research is needed to clarify their added value?

*Mark only one oval.*

- Yes
- No

43. **Q21: chronic illness** *

Do you currently suffer from one of these conditions? (multiple answer possible)

- [ ] Cancer
- [ ] Diabetes Mellitus
- [ ] High Blood Pressure
- [ ] Heart attack/coronary heart disease
- [ ] Congestive Heart Failure
- [ ] Cerebrovascular Disease
- [ ] Arthritis
- [ ] Chronic Lung Disease
- [ ] Stomach or Intestinal Ulcers
- [ ] Migraine
- [ ] Cataract
- [ ] Glaucoma
- [ ] None of the above

*Mark only one oval per row.*

| | Low | Medium | High |
|---|---|---|---|
| **Importance** | | | |
| **Burden** | | | |
| **Accuracy** | | | |

44. **Q21: chronic illness** *

Do you believe that this item is underexplored, and more research is needed to clarify their added value?

*Mark only one oval.*

- Yes
- No

45. **Q22: frailty index** *

[This index is a composite measure derived based on uses the EQ-5D (5 questions, reported below) and the SF36 questionnaire (36 questions, see online). Other measures could be potentially used to characterize the frailty level, e.g the Groningen Frailty Indicator (GFI).]

EQ-5D: For each group listed below, please put a cross behind the sentence that best reflects your health status today (only 1 cross per group). It is best to read all possible answers before you tick one.

Mobility
☐ I have no problems walking around
☐ I have some problems walking around
☐ I am confined to bed

Self-care
☐ I have no problems taking care of myself
☐ I have some problems washing or dressing myself
☐ I am unable to wash or dress myself

Daily activities (e.g. work, study, household, family or leisure activities)
☐ I have no problems with my daily activities
☐ I have some problems with my daily activities
☐ I am unable to perform my daily activities

Pain/discomfort
☐ I have no pain or discomfort
☐ I have moderate pain or discomfort
☐ I have very severe pain or discomfort

Anxiety/depression
☐ I am not anxious or depressed
☐ I am moderately anxious or depressed
☐ I am very anxious or depressed

*Mark only one oval per row.*

| | Low | Medium | High |
|---|---|---|---|
| **Importance** | ◯ | ◯ | ◯ |
| **Burden** | ◯ | ◯ | ◯ |
| **Accuracy** | ◯ | ◯ | ◯ |

46. **Q22: frailty index** *

Do you believe that this item is underexplored, and more research is needed to clarify their added value?

*Mark only one oval.*

- Yes
- No

47. **Q23: receiving care** *

Do you receive personal/medical/household care at home by family or professional caregivers?

- ☐ Yes, family outside household
- ☐ Yes, family inside household
- ☐ Yes, professional caregivers
- ☐ No

*Mark only one oval per row.*

| | Low | Medium | High |
|---|---|---|---|
| **Importance** | | | |
| **Burden** | | | |
| **Accuracy** | | | |

48. **Q23: receiving care** *

Do you believe that this item is underexplored, and more research is needed to clarify their added value?

*Mark only one oval.*

- Yes
- No

#### Other questions

If you want to review the instructions or the definition of the evaluation metrics, you can find them here.

49. **Q24: remote working / distance learning** *
How often do you currently work/study from home? Please indicate how many days per week.
[insert number]

*Mark only one oval per row.*

| | Low | Medium | High |
|---|---|---|---|
| **Importance** | ⬭ | ⬭ | ⬭ |
| **Burden** | ⬭ | ⬭ | ⬭ |
| **Accuracy** | ⬭ | ⬭ | ⬭ |

50. **Q24: remote working / distance learning** *
Do you believe that this item is underexplored, and more research is needed to clarify their added value?

*Mark only one oval.*

- Yes
- No

51. **Q25: change of behavior due to illness** *
You plan to dine in a restaurant, but you develop a fever and cough. Would you still go out to dine?
☐ Yes
☐ No

You plan to go shopping or to buy groceries from a market, but you develop a fever and cough. Would you still go out for shopping/grocery-buying?
☐ Yes
☐ No

Once you developed a fever and cough, how long would you wait to see the doctor or go to hospital? (days)
[insert number]

If you felt well, would you wear a face mask on public transport, in shopping centres, or markets?
☐ Yes
☐ No

*Mark only one oval per row.*

| | Low | Medium | High |
|---|---|---|---|
| **Importance** | ⬭ | ⬭ | ⬭ |
| **Burden** | ⬭ | ⬭ | ⬭ |
| **Accuracy** | ⬭ | ⬭ | ⬭ |

52. **Q25: change of behavior due to illness** *

Do you believe that this item is underexplored, and more research is needed to clarify their added value?

*Mark only one oval.*

- Yes
- No

53. **Q26: risk perception** *

To what extent do you agree or disagree with each of the following statements?

1. Coronavirus would be a serious illness for me
☐ Strongly agree ☐ Tend to agree ☐ Neither agree nor disagree ☐ Tend to disagree ☐ Strongly disagree ☐ Don't know

2. I am likely to catch coronavirus
☐ Strongly agree ☐ Tend to agree ☐ Neither agree nor disagree ☐ Tend to disagree ☐ Strongly disagree ☐ Don't know

3. If I don't follow the government's advice, I might spread coronavirus to someone who is vulnerable
☐ Strongly agree ☐ Tend to agree ☐ Neither agree nor disagree ☐ Tend to disagree ☐ Strongly disagree ☐ Don't know

*Mark only one oval per row.*

| | Low | Medium | High |
|---|---|---|---|
| **Importance** | | | |
| **Burden** | | | |
| **Accuracy** | | | |

54. **Q26: risk perception** *

Do you believe that this item is underexplored, and more research is needed to clarify their added value?

*Mark only one oval.*

- Yes
- No

55. **Q27: COVID-19 vaccination** *

Did you receive a COVID-19 vaccine in the last year/in the last winter campaign?

- [ ] Yes
- [ ] No
- [ ] Prefer not to answer

*Mark only one oval per row.*

| | Low | Medium | High |
|---|---|---|---|
| **Importance** | ◯ | ◯ | ◯ |
| **Burden** | ◯ | ◯ | ◯ |
| **Accuracy** | ◯ | ◯ | ◯ |

56. **Q27: COVID-19 vaccination** *

Do you believe that this item is underexplored, and more research is needed to clarify their added value?

*Mark only one oval.*

◯ Yes

◯ No

57. **Q28: influenza vaccination** *

Did you receive a flu vaccine in the last year/last winter campaign?

- [ ] Yes
- [ ] No
- [ ] Prefer not to answer

*Mark only one oval per row.*

| | Low | Medium | High |
|---|---|---|---|
| **Importance** | ◯ | ◯ | ◯ |
| **Burden** | ◯ | ◯ | ◯ |
| **Accuracy** | ◯ | ◯ | ◯ |

58. **Q28: influenza vaccination** *

Do you believe that this item is underexplored, and more research is needed to clarify their added value?

*Mark only one oval.*

◯ Yes

◯ No

#### SECTION 2: contact diary

This section contains questions designed to capture the characteristics of contacts reported by participants.

**Contact definition**

The contact definition used in this survey is taken from Willem *et al.* (PLOS One, 2012). A contact is defined as follows,

1. that you have spoken to someone in his/her presence at a distance of less than three meters (telephone contact or contact via the internet does not count)
2. or, that you have had physical contact with someone: if you have touched someone (even if it was without having a conversation), this is also a contact (e.g. shaking someone's hand, giving them a kiss, hugging, casual contacts during sports, etc.).

#### Contact features

**Important:** The following questions are asked once for **each contact** reported by the participant. As a result, the overall burden depends on the total number of contacts reported. For this survey, however, the focus is on the **burden per individual item**, not on cumulative burden. Please evaluate each question as if it were asked for **a single contact**, without considering repetition across contacts.

If you want to review the instructions or the definition of the evaluation metrics, you can find them here.

59. **Q29: age of contact** *

[ASK for each contact]

What is the age of this person?

☐☐ (– ☐☐ )

*Note: numbers in brackets can be used to indicate an age band when exact age is not known*

*Mark only one oval per row.*

| | Low | Medium | High |
|---|---|---|---|
| **Importance** | ◯ | ◯ | ◯ |
| **Burden** | ◯ | ◯ | ◯ |
| **Accuracy** | ◯ | ◯ | ◯ |

60. **Q29: age of contact** *

Do you believe that this item is underexplored, and more research is needed to clarify their added value?

*Mark only one oval.*

◯ Yes

◯ No

61. **Q30: sex of contact** *
[ASK for each contact]

What is the sex of this person?
☐ female
☐ male
☐ prefer not to answer

*Mark only one oval per row.*

| | Low | Medium | High |
|---|---|---|---|
| **Importance** | ◯ | ◯ | ◯ |
| **Burden** | ◯ | ◯ | ◯ |
| **Accuracy** | ◯ | ◯ | ◯ |

62. **Q30: sex of contact** *
Do you believe that this item is underexplored, and more research is needed to clarify their added value?

*Mark only one oval.*

◯ Yes

◯ No

63. **Q31: physical contact** *
[ASK for each contact]

Did you touch this person?
☐ Yes
☐ No

*Mark only one oval per row.*

| | Low | Medium | High |
|---|---|---|---|
| **Importance** | ◯ | ◯ | ◯ |
| **Burden** | ◯ | ◯ | ◯ |
| **Accuracy** | ◯ | ◯ | ◯ |

64. **Q31: physical contact** *
Do you believe that this item is underexplored, and more research is needed to clarify their added value?

*Mark only one oval.*

- Yes
- No

65. **Q32: contact frequency** *
[ASK for each contact]

How often do you meet this person?
☐ Daily or almost daily
☐ About once or twice a week
☐ About once or twice a month
☐ Less than once a month
☐ Never met before

*Mark only one oval per row.*

| | Low | Medium | High |
|---|---|---|---|
| **Importance** | | | |
| **Burden** | | | |
| **Accuracy** | | | |

66. **Q32: contact frequency** *
Do you believe that this item is underexplored, and more research is needed to clarify their added value?

*Mark only one oval.*

- Yes
- No

67. **Q33: location of contact** *
[ASK for each contact]

Where did you have contact?
(multiple answers possible)
☐ home
☐ work
☐ school
☐ transports
☐ leisure
☐ other

*Mark only one oval per row.*

| | Low | Medium | High |
|---|---|---|---|
| **Importance** | ◯ | ◯ | ◯ |
| **Burden** | ◯ | ◯ | ◯ |
| **Accuracy** | ◯ | ◯ | ◯ |

68. **Q33: location of contact** *
Do you believe that this item is underexplored, and more research is needed to clarify their added value?

*Mark only one oval.*

◯ Yes

◯ No

69. **The previous question (Q33)** includes six locations commonly used in social contact studies (home, work, school, transport, leisure, other). Some studies have included additional options to better characterize the *"other"* category (e.g., Kiti et al., *Emerging Infectious Diseases*, 2025; Latsuzbaia et al., *PLOS One*, 2020). *
Below, we propose some additional location options.

**Which of the following do you think would be useful to include as additional options for the location of the contact?**
*(Multiple answers possible)*

*Tick all that apply.*

☐ none
☐ others' home
☐ market/shop
☐ restaurant/bar/café
☐ sports
☐ place of worship
☐ Other: ____________________

70. **Q34: contact duration** *
[ASK for each contact]

How much time did you spend with this person throughout the day?
☐ under 5 minutes
☐ 5 — 15min
☐ 15min — 1h
☐ 1 — 4h
☐ more than 4h

*Mark only one oval per row.*

| | Low | Medium | High |
|---|---|---|---|
| **Importance** | ◯ | ◯ | ◯ |
| **Burden** | ◯ | ◯ | ◯ |
| **Accuracy** | ◯ | ◯ | ◯ |

71. **Q34: contact duration** *
Do you believe that this item is underexplored, and more research is needed to clarify their added value?

*Mark only one oval.*

◯ Yes

◯ No

72. **Q35: contact with household member** *
[ASK for each contact]

Is this person a member of your household?
☐ Yes
☐ No

*Mark only one oval per row.*

| | Low | Medium | High |
|---|---|---|---|
| **Importance** | ◯ | ◯ | ◯ |
| **Burden** | ◯ | ◯ | ◯ |
| **Accuracy** | ◯ | ◯ | ◯ |

73. **Q35: contact with household member** *

Do you believe that this item is underexplored, and more research is needed to clarify their added value?

*Mark only one oval.*

- Yes
- No

74. **Q36: relationship to this contact** *

[ASK for each contact]

What is this person's relationship to you?
Select only one
☐ household member
☐ family member who is not in my household
☐ someone I work with
☐ someone I go to school, college or university with
☐ friend
☐ other
☐ prefer not to answer

*Mark only one oval per row.*

| | Low | Medium | High |
|---|---|---|---|
| **Importance** | | | |
| **Burden** | | | |
| **Accuracy** | | | |

75. **Q36: relationship to this contact** *

Do you believe that this item is underexplored, and more research is needed to clarify their added value?

*Mark only one oval.*

- Yes
- No

76. **Q37: mask wearing** *
[ASK for each contact]

Did you wear a mask the entire time you were with this person?
☐ Yes
☐ No

*Mark only one oval per row.*

| | Low | Medium | High |
|---|---|---|---|
| **Importance** | ⬭ | ⬭ | ⬭ |
| **Burden** | ⬭ | ⬭ | ⬭ |
| **Accuracy** | ⬭ | ⬭ | ⬭ |

77. **Q37: mask wearing** *
Do you believe that this item is underexplored, and more research is needed to clarify their added value?

*Mark only one oval.*

⬭ Yes

⬭ No

78. **Q38: outdoor/indoor** *
[ASK for each contact]

Was the time that you spent with this person outside and/or inside?
☐ Inside
☐ Outside

*Mark only one oval per row.*

| | Low | Medium | High |
|---|---|---|---|
| **Importance** | ⬭ | ⬭ | ⬭ |
| **Burden** | ⬭ | ⬭ | ⬭ |
| **Accuracy** | ⬭ | ⬭ | ⬭ |

79. **Q38: outdoor/indoor** *
Do you believe that this item is underexplored, and more research is needed to clarify their added value?

*Mark only one oval.*

⬭ Yes

⬭ No

80. **Q39: contact in common** *

[ASK for each contact]

Did anyone that you live with also meet this person yesterday?
- ☐ Yes
- ☐ No
- ☐ Not sure

*Mark only one oval per row.*

| | Low | Medium | High |
|---|---|---|---|
| **Importance** | ◯ | ◯ | ◯ |
| **Burden** | ◯ | ◯ | ◯ |
| **Accuracy** | ◯ | ◯ | ◯ |

81. **Q39: contact in common** *

Do you believe that this item is underexplored, and more research is needed to clarify their added value?

*Mark only one oval.*

◯ Yes

◯ No

82. **Q40: ethnicity of contact** *

[ASK for each contact]

What is the ethnicity of this person?
- ☐ White
- ☐ Black
- ☐ Asian
- ☐ Hispanic / Latino
- ☐ Other
- ☐ Don't know

*Mark only one oval per row.*

| | Low | Medium | High |
|---|---|---|---|
| **Importance** | ◯ | ◯ | ◯ |
| **Burden** | ◯ | ◯ | ◯ |
| **Accuracy** | ◯ | ◯ | ◯ |

83. **Q40: ethnicity of contact** *

Do you believe that this item is underexplored, and more research is needed to clarify their added value?

*Mark only one oval.*

- Yes
- No

84. **Q41: time of contact** *

[ASK for each contact]

When did the contact occur?

- ☐ 5:00-8:00
- ☐ 8:00-12:00
- ☐ 12:00-18:00
- ☐ 18:00-21:00
- ☐ 21:00-5:00

*Mark only one oval per row.*

| | Low | Medium | High |
|---|---|---|---|
| **Importance** | | | |
| **Burden** | | | |
| **Accuracy** | | | |

85. **Q41: time of contact** *

Do you believe that this item is underexplored, and more research is needed to clarify their added value?

*Mark only one oval.*

- Yes
- No

#### Additional contacts

If you want to review the instructions or the definition of the evaluation metrics, you can find them here.

86. **Q42: additional contacts** *
We do ask you to individually include every contact you had between 5am yesterday and 5am today, but if you were unable to include every single contact (for instance, because you work in a shop and have a large number of contacts in a day), please could you indicate this?
☐ I individually included every person I had contact with.
☐ I did not individually include every person I had contact with.

*Mark only one oval per row.*

| | Low | Medium | High |
|---|---|---|---|
| **Importance** | ◯ | ◯ | ◯ |
| **Burden** | ◯ | ◯ | ◯ |
| **Accuracy** | ◯ | ◯ | ◯ |

87. **Q42: additional contacts** *
Do you believe that this item is underexplored, and more research is needed to clarify their added value?

*Mark only one oval.*

◯ Yes

◯ No

88. **Q42a: additional physical contacts** *
If you did not include every person you had contact with, approximately how many people you left out did you have
physical contact with: [insert number]
no physical contact with: [insert number]

*Mark only one oval per row.*

| | Low | Medium | High |
|---|---|---|---|
| **Importance** | ◯ | ◯ | ◯ |
| **Burden** | ◯ | ◯ | ◯ |
| **Accuracy** | ◯ | ◯ | ◯ |

89. **Q42a: additional physical contacts** *
Do you believe that this item is underexplored, and more research is needed to clarify their added value?

*Mark only one oval.*

◯ Yes

◯ No

90. **Q42b: additional contacts by setting** *
Approximately how many people did you have contact with and you did not list individually? Please provide your best estimate by setting.

At work: [insert number]
At school or other educational setting: [insert number]
Somewhere else: [insert number]

*Mark only one oval per row.*

| | Low | Medium | High |
|---|---|---|---|
| **Importance** | ◯ | ◯ | ◯ |
| **Burden** | ◯ | ◯ | ◯ |
| **Accuracy** | ◯ | ◯ | ◯ |

91. **Q42b: additional contacts by setting** *
Do you believe that this item is underexplored, and more research is needed to clarify their added value?

*Mark only one oval.*

◯ Yes

◯ No

92. **Q42c: additional contacts by age** *
[ASK for each LOCATION = work, educational setting, other settings]

Approximately how many people did you have contact with at LOCATION and you did not list individually? Please provide your best estimate by age.

Under 18: [insert number]
18 – 64: [insert number]
65 and over: [insert number]

*Mark only one oval per row.*

| | Low | Medium | High |
|---|---|---|---|
| **Importance** | ◯ | ◯ | ◯ |
| **Burden** | ◯ | ◯ | ◯ |
| **Accuracy** | ◯ | ◯ | ◯ |

93. **Q42c: additional contacts by age** *

Do you believe that this item is underexplored, and more research is needed to clarify their added value?

*Mark only one oval.*

- Yes
- No

94. **Q42d: duration of additional contacts** *

[ASK for each LOCATION = work, educational setting, other settings]

How much time did you spend with each person on average of the people you met at LOCATION?

- ☐ Less than 1 minute
- ☐ 1 minutes or more, but less than 5 minutes
- ☐ 5 minutes or more, but less than 15 minutes
- ☐ 15 minutes or more, but less than 1 hour
- ☐ 1 hour or more

*Mark only one oval per row.*

| | Low | Medium | High |
|---|---|---|---|
| **Importance** | | | |
| **Burden** | | | |
| **Accuracy** | | | |

95. **Q42d: duration of additional contacts** *

Do you believe that this item is underexplored, and more research is needed to clarify their added value?

*Mark only one oval.*

- Yes
- No

#### SECTION 3: information on the survey day

This is the last section of the questionnaire. This section contains questions about participants' behavior during the survey day.

#### Information about the survey day

If you want to review the instructions or the definition of the evaluation metrics, you can find them here.

96. **Q43: recall** *

How well do you recall your contacts?

- ☐ Very well
- ☐ Well
- ☐ Moderate well
- ☐ Not well
- ☐ Poorly

*Mark only one oval per row.*

| | Low | Medium | High |
|---|---|---|---|
| **Importance** | ⬭ | ⬭ | ⬭ |
| **Burden** | ⬭ | ⬭ | ⬭ |
| **Accuracy** | ⬭ | ⬭ | ⬭ |

97. **Q43: recall** *

Do you believe that this item is underexplored, and more research is needed to clarify their added value?

*Mark only one oval.*

⬭ Yes

⬭ No

98. **Q44: public transport** *

Did you use public transport on the assigned day?

- ☐ Yes, tram or bus
- ☐ Yes, train
- ☐ Yes, other
- ☐ No

*Mark only one oval per row.*

| | Low | Medium | High |
|---|---|---|---|
| **Importance** | ⬭ | ⬭ | ⬭ |
| **Burden** | ⬭ | ⬭ | ⬭ |
| **Accuracy** | ⬭ | ⬭ | ⬭ |

99. **Q44: public transport** *
Do you believe that this item is underexplored, and more research is needed to clarify their added value?

*Mark only one oval.*

- Yes
- No

100. **Q45: time use - location** *
[ASK for each time block]
5-8am, 8-10am, ..., 10-12pm,12pm-2am, 2-5am

For each time block for the day allotted to you, please indicate where you spent most of your time:
☐ home
☐ work
☐ school
☐ transports
☐ leisure
☐ other

*Mark only one oval per row.*

| | Low | Medium | High |
|---|---|---|---|
| **Importance** | | | |
| **Burden** | | | |
| **Accuracy** | | | |

101. **Q45: time use - location** *
Do you believe that this item is underexplored, and more research is needed to clarify their added value?

*Mark only one oval.*

- Yes
- No

102. **Q46: time use - distance from home** *

[ASK for each time block]
5-8am, 8-10am, ..., 10-12pm,12pm-2am, 2-5am

For each time block for the day allotted to you, please indicate how far from home you were during that time:
☐ 0-1 km
☐ 2-9km
☐ 10-74km
☐ 75km or more

*Mark only one oval per row.*

| | Low | Medium | High |
|---|---|---|---|
| **Importance** | ◯ | ◯ | ◯ |
| **Burden** | ◯ | ◯ | ◯ |
| **Accuracy** | ◯ | ◯ | ◯ |

103. **Q46: time use - distance from home** *

Do you believe that this item is underexplored, and more research is needed to clarify their added value?

*Mark only one oval.*

◯ Yes

◯ No

104. **Q47: crowding on transport** *

Make a list of all the transport you used on the day (taxi, private car, bus...).

[ASK for each trip]
How many people were on the vehicle at the start of your trip?
[insert number]

*Mark only one oval per row.*

| | Low | Medium | High |
|---|---|---|---|
| **Importance** | ◯ | ◯ | ◯ |
| **Burden** | ◯ | ◯ | ◯ |
| **Accuracy** | ◯ | ◯ | ◯ |

105. **Q47: crowding on transport** *

Do you believe that this item is underexplored, and more research is needed to clarify their added value?

*Mark only one oval.*

- Yes
- No

106. **Q48: crowding in indoor locations** *

Make a list of all the indoor places you visited on the survey day.
[ASK for each place]
What type of location was it?
Example of possible answers: Church, Clinic, Gym, Library, Mall / Shops, Bar/ Nightclub, School, Office, Restaurant, Other

Is this place your workplace? Yes/No

How long did you spend there? [insert duration]

How many people were there, halfway through the time you were there? [insert number]

*Mark only one oval per row.*

| | Low | Medium | High |
|---|---|---|---|
| **Importance** | | | |
| **Burden** | | | |
| **Accuracy** | | | |

107. **Q48: crowding in indoor locations** *

Do you believe that this item is underexplored, and more research is needed to clarify their added value?

*Mark only one oval.*

- Yes
- No

108. **Q49: working / vacation** *

Did you work on the day of the survey?

- [ ] Yes, I was at my workplace
- [ ] Yes, I worked from home
- [ ] No, I had a day off
- [ ] No, I was sick

*Mark only one oval per row.*

| | Low | Medium | High |
|---|---|---|---|
| **Importance** | ◯ | ◯ | ◯ |
| **Burden** | ◯ | ◯ | ◯ |
| **Accuracy** | ◯ | ◯ | ◯ |

109. **Q49: working / vacation** *

Do you believe that this item is underexplored, and more research is needed to clarify their added value?

*Mark only one oval.*

◯ Yes

◯ No

110. **Q50: symptoms on survey day** *

Did you have any of the following symptoms on the day of the survey?

- [ ] Fever or high temperature
- [ ] A cough that has lasted for at least several hours
- [ ] Shortness of breath
- [ ] Aches and pains, e.g. in back, neck, shoulders or joints
- [ ] Blocked nose
- [ ] Sore throat
- [ ] Feeling unusually tired
- [ ] No symptoms
- [ ] Don’t know
- [ ] Prefer not to answer

*Mark only one oval per row.*

| | Low | Medium | High |
|---|---|---|---|
| **Importance** | ◯ | ◯ | ◯ |
| **Burden** | ◯ | ◯ | ◯ |
| **Accuracy** | ◯ | ◯ | ◯ |

111. **Q50: symptoms on survey day** *
Do you believe that this item is underexplored, and more research is needed to clarify their added value?

*Mark only one oval.*

- Yes
- No

*Skip to question 112*

#### Thank you.

112. Thank you for taking the time to complete this survey. If you have any additional comments or suggestions related to the content, wording, or structure to improve social contact data collection, please share them below.

This content is neither created nor endorsed by Google.

Google Forms
