## Supplementary Material 6 for "On conducting diary-based social contact surveys to inform transmission models of close-contact infections: a systematic review and expert-informed questionnaire"

Overview of survey items from studies conducted in general community settings and their classification. References indicate studies in which the corresponding survey item was explicitly asked in the questionnaire. Studies included in the systematic review that did not report the survey items explicitly are not referenced here.

| Classification | Category | Survey items | Question examples | References |
|---|---|---|---|---|
| **Core (n = 22)** | Demographic factors | Age | What is your age? | [1–36] |
| | | Place of residence | What is the postal code of your place of residence? | [1–5,7–10,13–15,17,19,20,22–24,26–32,35] |
| | | Sex | What is your sex? | [2–5,7–10,12–36] |
| | Socio-economic factors | Education level | What is the highest level of education you have completed? | [3,8–10,13,16,17,19,23,24,26–30,34–36] |
| | | Employment status | What is your current employment status? | [2–4,6,8–10,13,17–19,21,22,24–30,36] |
| | | Occupation | What is your occupation? | [1,3,7,8,10,12,13,15,16,18–21,23,24,26,27] |
| | Household composition | Age of household members | [Ask for each household member] What is their age? | [1–4,8–10,12,13,15,18–21,24,26,27,29,32] |
| | | Household size | Including you, how many other people live in your household? | [1,2,4–13,15,16,18–29,31–36] |
| | Health status | Health-seeking behaviour | 1. Did you (or anyone else in your household) have any of the following symptoms in the last 7 days?<br>2. Have you (or anyone else in your household) sought any medical services following these symptoms? | [1,5,18,20,21,23,26,27] |
| | Other (participant) questions | Change behaviour due to illness | 1. You plan to dine in a restaurant, but you develop a fever and cough. Would you still go out to dine?<br>2. You plan to go shopping or to buy groceries from a market, but you develop a fever and cough. Would you still go?<br>3. Once you developed a fever and cough, how long would you wait to see the doctor or go to the hospital (days)?<br>4. If you felt well, would you wear a face mask on public transport, in shopping centres or markets? | [1,2,4–6,25] |
| | | Remote working status | How often do you currently work/study from home? Please indicate how many days per week. | [1,2,19] |
| | Contact features | Age of contact | [Ask for each contact] What is the age of this person? | [1–10,12,13,15,16,16–24,29–36] |
| | | Sex of contacts | [Ask for each contact] What is the sex of this person? | [7,8,12,13,16,18,20–24,26,27,29–36] |
| | | Contact duration | [Ask for each contact] How much time did you spend with this person throughout the day? | [7–9,12,13,15,16,18,20–24,26,27,29–33,33–36] |
| | | Contact frequency | [Ask for each contact] How often do you meet this person? | [7,9,12,13,15,16,18,20–22,24,26,29–33,36] |
| | | Contact with household members | [Ask for each contact] Is this person a member of your household? | [5,6,9,23,26,27,30,31,33,36] |
| | | Outdoor contacts | [Ask for each contact] Was the time that you spent with this person outside and/or inside? | [3,4,9,13,18,21,26,27,30,31] |
| | | Physical contacts | [Ask for each contact] Did you touch this person? | [2,3,6–10,12,13,15–24,26,27,30–32,34–36] |
| | | Location of contacts | [Ask for each contact] Where did you have contact? | [1–3,5,7–9,11–13,15–21,23–26,28–32,34–36] |
| | Information about the survey day | Symptoms on the survey day | Did you have any of the following symptoms on the day of the survey? | [13,14,33] |
| | | Working vacation status | Did you work on the day of the survey? | [10] |
| | Additional contacts | Additional contacts | We do ask you to individually include every contact you had between 5 am yesterday and 5 am today, but if you were unable to include every single contact (for instance, because you work in a shop and have a large number of contacts in a day), please could you indicate this? | [18,26,27] |

| Classification | Category | Survey items | Question examples | References |
|---|---|---|---|---|
| **Secondary (n = 14)** | Demographic factors | Ethnicity | Which ethnic group do you belong to? | [1,3–5,22,23,26,27,29,29,32] |
| | Socio-economic factors | Individual/household income | Into which category does your total net (household) monthly income from all sources fall (i.e. income after tax deduction)?<br>Please include all your (household) income sources: salaries, scholarships, pension and Social Security benefits, dividends from shares, income from rental properties, child support and alimony, etc. | [1,3,5,8,18,21–24,26,27,36] |
| | Household composition | Household type | Which of the following best describes your household? | [2–5,8,10,13,18,20–23,26,27] |
| | | Vaccination status of household members | [Ask for each household member] Did they receive XXX vaccine in the last year/in the last winter campaign? | [1,26,27] |
| | Health status | Chronic illness | Do you currently suffer from one of these (medical) conditions? (multiple answers possible) | [5,10,13,16,30] |
| | Other (participant) questions | Influenza vaccination | Did you receive a flu vaccine in the last year/last winter campaign? | [17,23] |
| | | COVID-19 vaccination | Did you receive a COVID-19 vaccine in the last year/in the last winter campaign? | [13,17,26,27] |
| | | Risk perception | To what extent do you agree or disagree with each of the following statements?<br><br>1. Coronavirus would be a serious illness for me<br>2. I am likely to catch coronavirus<br>3. If I don't follow the government's advice, I might spread coronavirus to someone vulnerable | [1,2,4,5,18,21,26–28] |
| | Contact features | Relationship to contactee | [Ask for each contact] What is this person's relationship to you? | [3,7,12,13,15,16,16,18,21,24,26,27,29,31,32] |
| | | Mask wearing | Did you wear a mask the entire time you were with this person? | [2,4,9,11,13,26,27,34–36] |
| | Information about the survey day | Public transport | Did you use public transport on the assigned day? | [4,9,10,13,16,18,21,26,27,31] |
| | Additional contacts | Additional contacts by setting | Approximately how many people did you have contact with that you did not list individually? Please provide your best estimate by setting.<br><br>At work: [insert number]<br>At school or other educational setting: [insert number]<br>Somewhere else: [insert number] | [18,26,27] |
| | | Additional contacts by age | [ASK for each LOCATION = work, educational setting, other settings]<br><br>Approximately how many people did you have contact with at LOCATION and you did not list individually? Please provide your best estimate by age.<br><br>Under 18: [insert number]<br>18 – 64: [insert number]<br>65 and over: [insert number] | [18,26,27] |
| | | Additional contacts by physical | If you did not include every person you had contact with, approximately how many people did you leave out<br><br>Physical contact with: [insert number]<br>No physical contact with: [insert number] | [18,26,27] |

| Classification | Category | Survey items | Question examples | References |
|---|---|---|---|---|
| **Optional (n = 18)** | Demographic factors | Country of birth | In which country were you born? | [11,13,28,29] |
| | | Country of birth of parents or grandparents | In which country were your parents/grandparents born? | [13] |
| | Household composition | Household structure | 1. What is the total number of rooms in your house?<br>2. How many rooms are used to sleep in your house? | [5,6,9,12,16,20] |
| | | Employment status of household members | [Ask for each household member] What is their current employment status? | [5,9,18,21,26,27,33] |
| | | Relationship of household members | [Ask for each household member] How is this member related to you? | [9,12,32] |
| | | Sex of household members | [Ask for each household member] What is their sex? | [8,9,12,13,18,20,21,24,26,27,29,32] |
| | Health status | Receiving care | Do you receive care at home by family or professional caregivers? | [13] |
| | | Pregnant status | Are you currently pregnant? | [18,21,23,26,27] |
| | | Frailty index | This index is a composite measure derived from e.g., the EQ-5D (5 questions), the SF36 questionnaire (36 questions), or the Groningen Frailty Indicator (GFI). | [13] |
| | Contact features | Time of contact | [Ask for each contact] When did the contact occur? | [23,24] |
| | | Shared contacts with household members | [Ask for each contact] Did anyone that you live with also meet this person yesterday? | [4] |
| | | Ethnicity of contact | What is the ethnicity of this person? | [29] |
| | Information about the survey day | Distance from home | [Ask for each time block] For each time block for the day allotted to you, please indicate how far from home you were during that time. | [4,13,16] |
| | | Time use location | [Ask for each time block] For each time block for the day allotted to you, please indicate where you spent most of your time. | [1,9,12,13,30–32] |
| | | Crowding on transport | Make a list of all the transportation you used on the day.<br>For each trip, how many people were on the vehicle at the start of your trip? | [31] |
| | | Crowding indoor | Make a list of all the indoor places you visited on the survey day.<br>1. What type of location was it?<br>2. Is this place your workplace?<br>3. How long did you spend there?<br>4. How many people were there halfway through the time you were there? | [9] |
| | | Recall | How well do you recall your contacts? | [23,24] |
| | Additional contacts | Additional contacts by duration | [Ask for each LOCATION] How much time did you spend with each person on average of the people you met at LOCATION? | [18,26,27] |
