## Supplementary Material 7 for "On conducting diary-based social contact surveys to inform transmission models of close-contact infections: a systematic review and expert-informed questionnaire"

This supplementary material presents the expert-informed contact questionnaire developed from the systematic review and structured expert assessment. The questionnaire comprises the 22 core items, together with instructions for completion and examples to support consistent interpretation and reporting. Non-core items are not included in this version of the questionnaire and are provided separately in Supplementary material 6.

The full expert-informed contact questionnaire is presented on the following page.

### INSTRUCTIONS

Please read these instructions carefully before you start filling in the contact diary.

1. In this study, you will be asked **to complete two sets of questions**. The first is a short questionnaire about your **demographics and details regarding the reference day of the survey**. The second is a **contact diary**, in which you will record every person you had contact with on your assigned day.

2. A contact is any person with whom you had at least one of the following interactions:
   - a **two-way conversation** with a few words in the physical presence of another person (at a distance of less than three meters);
   - physical **skin-to-skin contact** (for example, a handshake, hug, kiss, or contact sports).

   Contacts made exclusively by phone or internet (such as phone calls, video calls or online meetings) should not be recorded.

3. For the purposes of this study, your **reference day** is the day for which you report your contacts. The reference day begins at 5:00 a.m. and ends at 5:00 a.m. the following morning. For example, if you are assigned Monday as your reference day, report all contacts made between 5:00 a.m. on Monday and 5:00 a.m. on Tuesday.

4. Write down **every person that you contact** during the day, regardless of whether the contact was long or short, and whether you know the person or not.

5. The **order** of entries **is not important.** It might be easiest to list contacts chronologically based on when you first met them during daily activities, but feel free to order differently

6. Each contacted person corresponds to one line in the diary: **one person, one line.** If you had contact with the same person multiple times within the assigned day, record that person **only once** and provide an estimate of the **total time spent together across the entire day**. If other aspects of the contact vary (e.g., location), you may select **multiple answers** to reflect the different situations in which you met the same person. **For each contact, you will be asked to report** information such as:

| **Variable** | **Description** |
|---|---|
| Age | Age of contact (exact or estimated range).<br><br>Note: If you do not know the exact age of the contact, provide your best estimate (e.g. 40 to 45 years). For babies younger than 1 year, report age 0 years. |
| Sex | Sex of the contact person, based on your best judgment if unknown. |
| Skin contact | Whether physical skin contact occurred.<br><br>Note: Any brief physical contact should be recorded as “Yes” in the skin contact question. |
| Location(s) | Where the contact occurred from your perspective (e.g., a teacher records classroom contacts as work; a student records the same as school). |
| Location setting | Specify if the contact occurred Indoors (e.g., inside a building, house, or vehicle) or Outdoors (e.g., in the open air).<br><br>Note: If the contact moved between both settings, please check both boxes. |
| Frequency of contact | How often you usually interact with this person. |
| Duration | Estimated total contact duration over the day (e.g., less than 5 minutes, 5-15 minutes, 15-60 minutes, 1-4 hours, more than 4 hours).<br><br>Note: If you contact the same person multiple times within a day, record them only once and indicate an estimate of the total duration spent with them |
| Household member | Whether the person you had contact with normally lives at your address and shares the kitchen.<br><br>Note: This includes children living in student accommodation who regularly return home on weekends and are still considered part of the household. |

7. Please note that some **location categories** (home, others home, work, school, restaurant/bar/cafe, leisure, transportation, others) are grouped for simplicity. For example, “**School**” refers to all educational institutions (including childcare, schools, and universities), “**Transport**” includes both private and public modes of transportation, and "**Others home**" refers to any private residence other than your own, such as a visit to a friend, family member, or colleague.

8. After completing the diary, we recommend **double-checking your entries** by recalling all of your activities to ensure no contacts were missed.

#### DEMOGRAPHIC DATA

Please take some time to fill in the following details about yourself

| | |
|---|---|
| 1. Date of birth (DD/MM/YYYY) | __.__/__.__/__.__.__.__ |
| 2. Sex | __ Male<br>__ Female<br>__ Other |
| 3. Postal code of your usual place of residence<br>*(i.e. where you spend most nights)* | __.__.__.__ |
| 4. Highest level of formal education completed<br>*(Select only one)* | __ No formal education / incomplete primary school<br>__ Primary education / no diploma<br>__ Lower secondary education<br>__ Upper secondary education<br>__ Non-degree education<br>__ Bachelor's degree<br>__ Postgraduate<br>__ Prefer not to say |
| 5. Employment status<br>*Please note: if more than one category applies, please select the one that best reflects the activity you spend the most time on in your daily life.* | __ Employed full-time (34 hours or more per week)<br>__ Employed part-time (less than 34 hours per week)<br>__ Self-employed / freelancer<br>__ Student (includes those working part-time)/pupil<br>__ Unemployed (and those looking for work)<br>__ Homemaker/caregiver<br>__ Retired (including early retirement)<br>__ Inactive (e.g. due to disability or long-term sickness)<br>__ Prefer not to say |
| 6. Primary occupation<br>*(Select only one)*<br>*Please note: if you are not currently working but have worked in the past (e.g. you are retired, temporarily unemployed or currently unable to work), please refer to your most recent occupation.* | __ Not applicable (never worked)<br>__ Professional/technical<br>__ Managerial/executive<br>__ Clerical/administrative support<br>__ Sales and service<br>__ Skilled trades/construction<br>__ Agriculture, forestry, and fishing<br>__ Manufacturing, logistics, and manual labour<br>__ Healthcare and social assistance<br>__ Others<br>__ Prefer not to say |
| 7. Number of people living in your household<br>*Please note: by household size, we mean the total number of people (including yourself) living at the same address as you, that you share a kitchen with. If you live in a collective household setting (e.g., student housing or a retirement home), please provide your best estimate.* | ______ |
| 8a. If you live in a private household (e.g., a house or apartment), please list the ages of all household members (including yourself), starting with the youngest.<br>*Please note: if you live in a collective household, please leave this section blank and proceed to Question 8b.* | _____<br>_____<br>_____<br>_____<br>_____<br>_____<br>_____<br>_____<br>_____ |
| 8b. If you live in a collective household (e.g., student housing, retirement home, or similar institution), please indicate the total number of residents living in the entire facility. | __ Less than 10 people<br>__ Between 10 and 29 people<br>__ Between 30 and 49 people<br>__ Between 50 and 99 people<br>__ More than 100 people |

#### REFERENCE DAY CHARACTERISTICS

Please take some time to fill in the following details about the reference day

| | |
|---|---|
| 9. In a typical week, on how many days do you do most of your work or study from home or another location outside your usual workplace or educational setting?<br><br>*Please include any day where most of your work or study activities are carried out remotely rather than at your usual workplace or educational setting.* | __ days per week |
| 10. During your reference day (5:00 am on your assigned day to 5:00 am the next morning), which of the following best describes your status? | __ I worked or studied in person (at a workplace, school, or university)<br>__ I mostly stayed at home (e.g., teleworking, distance learning, or home office)<br>__ I was on sick leave (and did not work/study)<br>__ I was on holiday or a scheduled day off<br>__ Other |

11. Please indicate if you have experienced any of the following symptoms during your reference day or at any time during the last 7 days.

| **Symptom** | **During your reference day** | **Experienced in the last 7 days** |
|---|---|---|
| Runny or blocked (stuffy) nose | __ | __ |
| Sneezing | __ | __ |
| Sore throat | __ | __ |
| Loss of smell or taste | __ | __ |
| Headache | __ | __ |
| Fever | __ | __ |
| Cold chills | __ | __ |
| Fatigue or malaise | __ | __ |
| Aches or muscle pain | __ | __ |
| Loss of appetite | __ | __ |
| Cough (dry or productive) | __ | __ |
| Difficulty breathing or shortness of breath | __ | __ |
| Chest pain | __ | __ |
| Nausea | __ | __ |
| Eye pain | __ | __ |
| Other symptoms not listed above | __ | __ |
| No symptoms | __ | __ |

| | |
|---|---|
| 12. If you reported any symptoms during your reference day or in the last 7 days, did you seek any of the following medical services for the symptoms? | __ Phoned medical emergency number<br>__ Phoned a GP practice/GP out of hours service<br>__ Visited a GP practice/GP out of hours service<br>__ Visited Accident & Emergency (A&E)<br>__ Visited a testing location somewhere different to these services<br>__ Been admitted to hospital<br>__ Don’t know<br>__ None of these<br>__ Prefer not to answer |

#### CONTACT DIARY: EXAMPLES

**Reference date:** ☐☐/☐☐/☐☐☐☐ (DD/MM/YYYY)

You are a 30-year-old male working as a high-school teacher. Below are examples of how you might record your contacts based on your daily activities.

1. In the morning, you drove your 9-year-old son to school and chatted with him for about 10 minutes in the car. Later in the afternoon, you spent 1 hour together gardening in your backyard. In the evening, you played from 6:00 to 9:00 p.m. and kissed him goodnight before bed. (direct in person conversation + skin-to-skin contact → **relevant**)
2. In the staff lounge, you had a 30-minute discussion with a fellow female teacher from another department whom you were meeting for the first time. (direct in-person conversation → **relevant**)
3. After your colleague left the staff lounge, you remained there during your break. One other person was present, seated more than 3 meters away (presence > 3 meters → **not relevant**)
4. In the afternoon, you stopped by a shoe store. You spoke with a young saleswoman while trying on several pairs of shoes. (direct in-person conversation → **relevant**)
5. Afterwards, you went to visit your 65-year-old mother at her home. You visited her several times a week and stayed for about two hours discussing your day. (direct in-person conversation → **relevant**)
6. In the evening, you had a video call with a friend who lives abroad. (online conversation → **not relevant**)

| **Age** | **Sex** | | **Skin touch** | | **Household member** | | **Location(s) of the contact(s)** | | | | | | | | **Location setting** | | **Total time spent with this person during the day** | | | | | **Usual frequency of contact with this person** | | | | |
|---|---|---|---|---|---|---|---|---|---|---|---|---|---|---|---|---|---|---|---|---|---|---|---|---|---|---|
| (exact or approximated range) | F | M | Y | N | Y | N | My home | School | Work | Transport | Leisure | Restaurant/bar/cafe | Other homes | Other | Indoor | Outdoor | <5min | 5-15min | 15min-1h | 1-4 h | >4 h | Daily or almost daily | Many times a week | Many times a month | Many times a year or less | First time |
| 9(- ☐☐) | ☐ | × | × | ☐ | × | ☐ | × | ☐ | ☐ | × | ☐ | ☐ | ☐ | ☐ | × | × | ☐ | ☐ | ☐ | × | ☐ | × | ☐ | ☐ | ☐ | ☐ |
| 30 (- 36) | × | ☐ | ☐ | × | ☐ | × | ☐ | ☐ | × | ☐ | ☐ | ☐ | ☐ | ☐ | × | ☐ | ☐ | ☐ | × | ☐ | ☐ | ☐ | ☐ | ☐ | ☐ | × |
| 25 (- 30) | × | ☐ | ☐ | × | ☐ | × | ☐ | ☐ | ☐ | ☐ | ☐ | ☐ | ☐ | × | × | ☐ | ☐ | ☐ | × | ☐ | ☐ | ☐ | ☐ | ☐ | ☐ | × |
| 65(- ☐☐) | × | ☐ | ☐ | × | ☐ | × | ☐ | ☐ | ☐ | ☐ | ☐ | ☐ | × | ☐ | × | ☐ | ☐ | ☐ | ☐ | × | ☐ | ☐ | × | ☐ | ☐ | ☐ |
| ☐☐(- ☐☐) | ☐ | ☐ | ☐ | ☐ | ☐ | ☐ | ☐ | ☐ | ☐ | ☐ | ☐ | ☐ | ☐ | ☐ | ☐ | ☐ | ☐ | ☐ | ☐ | ☐ | ☐ | ☐ | ☐ | ☐ | ☐ | ☐ |
| ☐☐(- ☐☐) | ☐ | ☐ | ☐ | ☐ | ☐ | ☐ | ☐ | ☐ | ☐ | ☐ | ☐ | ☐ | ☐ | ☐ | ☐ | ☐ | ☐ | ☐ | ☐ | ☐ | ☐ | ☐ | ☐ | ☐ | ☐ | ☐ |
| ☐☐(- ☐☐) | ☐ | ☐ | ☐ | ☐ | ☐ | ☐ | ☐ | ☐ | ☐ | ☐ | ☐ | ☐ | ☐ | ☐ | ☐ | ☐ | ☐ | ☐ | ☐ | ☐ | ☐ | ☐ | ☐ | ☐ | ☐ | ☐ |
| ☐☐(- ☐☐) | ☐ | ☐ | ☐ | ☐ | ☐ | ☐ | ☐ | ☐ | ☐ | ☐ | ☐ | ☐ | ☐ | ☐ | ☐ | ☐ | ☐ | ☐ | ☐ | ☐ | ☐ | ☐ | ☐ | ☐ | ☐ | ☐ |
| ☐☐(- ☐☐) | ☐ | ☐ | ☐ | ☐ | ☐ | ☐ | ☐ | ☐ | ☐ | ☐ | ☐ | ☐ | ☐ | ☐ | ☐ | ☐ | ☐ | ☐ | ☐ | ☐ | ☐ | ☐ | ☐ | ☐ | ☐ | ☐ |

### CONTACT DIARY

□□/□□/□□□□ (DD/MM/YYYY)

Please take a moment to fill in the following details about your contacts.

| Age | Sex | | Skin touch | | Household member | | Location(s) of the contact(s) | | | | | | | | Location setting | | Total time spent with this person during the day | | | | | Usual frequency of contact with this person | | | | |
|---|---|---|---|---|---|---|---|---|---|---|---|---|---|---|---|---|---|---|---|---|---|---|---|---|---|---|
| (exact or approximated range) | F | M | Y | N | Y | N | Home | School | Work | Transport | Leisure | Restaurant/bar/cafe | Other homes | Other | Indoor | Outdoor | <5min | 5-15min | 15min-1h | 1-4 h | >4 h | Daily or almost daily | Many times a week | Many times a month | Many times a year or less | First time |
| □□(- □□) | □ | □ | □ | □ | □ | □ | □ | □ | □ | □ | □ | □ | □ | □ | □ | □ | □ | □ | □ | □ | □ | □ | □ | □ | □ | □ |
| □□(- □□) | □ | □ | □ | □ | □ | □ | □ | □ | □ | □ | □ | □ | □ | □ | □ | □ | □ | □ | □ | □ | □ | □ | □ | □ | □ | □ |
| □□(- □□) | □ | □ | □ | □ | □ | □ | □ | □ | □ | □ | □ | □ | □ | □ | □ | □ | □ | □ | □ | □ | □ | □ | □ | □ | □ | □ |
| □□(- □□) | □ | □ | □ | □ | □ | □ | □ | □ | □ | □ | □ | □ | □ | □ | □ | □ | □ | □ | □ | □ | □ | □ | □ | □ | □ | □ |
| □□(- □□) | □ | □ | □ | □ | □ | □ | □ | □ | □ | □ | □ | □ | □ | □ | □ | □ | □ | □ | □ | □ | □ | □ | □ | □ | □ | □ |
| □□(- □□) | □ | □ | □ | □ | □ | □ | □ | □ | □ | □ | □ | □ | □ | □ | □ | □ | □ | □ | □ | □ | □ | □ | □ | □ | □ | □ |
| □□(- □□) | □ | □ | □ | □ | □ | □ | □ | □ | □ | □ | □ | □ | □ | □ | □ | □ | □ | □ | □ | □ | □ | □ | □ | □ | □ | □ |
| □□(- □□) | □ | □ | □ | □ | □ | □ | □ | □ | □ | □ | □ | □ | □ | □ | □ | □ | □ | □ | □ | □ | □ | □ | □ | □ | □ | □ |
| □□(- □□) | □ | □ | □ | □ | □ | □ | □ | □ | □ | □ | □ | □ | □ | □ | □ | □ | □ | □ | □ | □ | □ | □ | □ | □ | □ | □ |
| □□(- □□) | □ | □ | □ | □ | □ | □ | □ | □ | □ | □ | □ | □ | □ | □ | □ | □ | □ | □ | □ | □ | □ | □ | □ | □ | □ | □ |
| □□(- □□) | □ | □ | □ | □ | □ | □ | □ | □ | □ | □ | □ | □ | □ | □ | □ | □ | □ | □ | □ | □ | □ | □ | □ | □ | □ | □ |
| □□(- □□) | □ | □ | □ | □ | □ | □ | □ | □ | □ | □ | □ | □ | □ | □ | □ | □ | □ | □ | □ | □ | □ | □ | □ | □ | □ | □ |
| □□(- □□) | □ | □ | □ | □ | □ | □ | □ | □ | □ | □ | □ | □ | □ | □ | □ | □ | □ | □ | □ | □ | □ | □ | □ | □ | □ | □ |
| □□(- □□) | □ | □ | □ | □ | □ | □ | □ | □ | □ | □ | □ | □ | □ | □ | □ | □ | □ | □ | □ | □ | □ | □ | □ | □ | □ | □ |
| □□(- □□) | □ | □ | □ | □ | □ | □ | □ | □ | □ | □ | □ | □ | □ | □ | □ | □ | □ | □ | □ | □ | □ | □ | □ | □ | □ | □ |

The following questions ask about what you would usually do if you developed a fever and cough (for example, due to a respiratory infection). Please answer based on what you would most likely do.

1. If you developed a fever and cough, how likely would you be to go to your workplace or classroom?
   __ Very unlikely __ Unlikely __ Neutral __ Likely __ Very likely

2. If you planned to dine at a restaurant but developed a fever and cough, how likely would you be to still go?
   __ Very unlikely __ Unlikely __ Neutral __ Likely __ Very likely

3. If you planned to go shopping or buy groceries but developed a fever and cough, how likely would you be to still go?
   __ Very unlikely __ Unlikely __ Neutral __ Likely __ Very likely

4. If you felt well, how likely would you be to wear a face mask on public transport or in indoor public places (e.g. shopping centres or markets)?
   __ Very unlikely __ Unlikely __ Neutral __ Likely __ Very likely

5. If you developed a fever and cough, how soon would you usually seek medical care (e.g. visit a doctor or hospital)?
   __ Same day
   __ 1-2 days
   __ 3-5 days
   __ More than 5 days
   __ Only if symptoms worsen
   __ Would not seek care

We do ask you to include every contact you had, but if you were unable to include every single contact (e.g, because you work in a shop till and have a large number of contacts in a day), please could you indicate this?

__ I included every person I had contact with.
__ I did not include every person I had contact with.

If you did not include every person you had contact with, approximately how many people did you leave out
__________

Once again, thank you for your participation. All the information you have provided will be treated in strict confidence and will be used for research purposes only.
